# Multiomic foundations of human prefrontal cortex tissue function

**DOI:** 10.1101/2024.05.17.24307537

**Authors:** Brian H. Kopell, Deepak A. Kaji, Lora E. Liharska, Eric Vornholt, Alissa Valentine, Anina Lund, Alice Hashemi, Ryan C. Thompson, Terry Lohrenz, Jessica S. Johnson, Nicole Bussola, Esther Cheng, You Jeong Park, Punit Shah, Weiping Ma, Richard Searfoss, Salman Qasim, Gregory M. Miller, Nischal Mahaveer Chand, Alisha Aristel, Jack Humphrey, Lillian Wilkins, Kimia Ziafat, Hannah Silk, Lisa M. Linares, Brendan Sullivan, Claudia Feng, Seth R. Batten, Dan Bang, Leonardo S. Barbosa, Thomas Twomey, Jason P. White, Marina Vannucci, Beniamino Hadj-Amar, Vanessa Cohen, Prashant Kota, Emily Moya, Marysia-Kolbe Rieder, Martijn Figee, Girish N. Nadkarni, Michael S. Breen, Kenneth T. Kishida, Joseph Scarpa, Douglas M. Ruderfer, Niven R. Narain, Pei Wang, Michael A. Kiebish, Eric E. Schadt, Ignacio Saez, P. Read Montague, Noam D. Beckmann, Alexander W. Charney

**Author notes:** – Authors contributed equally (Brian H. Kopell, Deepak A. Kaji, Lora E. Liharska, Eric Vornholt). – Authors contributed equally (Michael A. Kiebish, Eric E. Schadt, Ignacio Saez, P. Read Montague). – Authors contributed equally (Noam D. Beckmann, Alexander W. Charney). Correspondence: Alexander W. Charney Noam D. Beckmann Brian H. Kopell.

## Abstract

The prefrontal cortex (PFC) is a region of the brain that in humans is involved in the production of higher-order functions such as cognition, emotion, perception, and behavior. Neurotransmission in the PFC produces higher-order functions by integrating information from other areas of the brain. At the foundation of neurotransmission, and by extension at the foundation of higher-order brain functions, are an untold number of coordinated molecular processes involving the DNA sequence variants in the genome, RNA transcripts in the transcriptome, and proteins in the proteome. These “multiomic” foundations are poorly understood in humans, perhaps in part because most modern studies that characterize the molecular state of the human PFC use tissue obtained when neurotransmission and higher-order brain functions have ceased (i.e., the postmortem state). Here, analyses are presented on data generated for the Living Brain Project (LBP) to investigate whether PFC tissue from individuals with intact higher-order brain function has characteristic multiomic foundations. Two complementary strategies were employed towards this end. The first strategy was to identify in PFC samples obtained from living study participants a signature of RNA transcript expression associated with neurotransmission measured intracranially at the time of PFC sampling, in some cases while participants performed a task engaging higher-order brain functions. The second strategy was to perform multiomic comparisons between PFC samples obtained from individuals with intact higher-order brain function at the time of sampling (i.e., living study participants) and PFC samples obtained in the postmortem state. RNA transcript expression within multiple PFC cell types was associated with fluctuations of dopaminergic, serotonergic, and/or noradrenergic neurotransmission in the substantia nigra measured while participants played a computer game that engaged higher-order brain functions. A subset of these associations – termed the “transcriptional program associated with neurotransmission” (TPAWN) – were reproduced in analyses of brain RNA transcript expression and intracranial neurotransmission data obtained from a second LBP cohort and from a cohort in an independent study. RNA transcripts involved in TPAWN were found to be (1) enriched for RNA transcripts associated with measures of neurotransmission in rodent and cell models, (2) enriched for RNA transcripts encoded by evolutionarily constrained genes, (3) depleted of RNA transcripts regulated by common DNA sequence variants, and (4) enriched for RNA transcripts implicated in higher-order brain functions by human population genetic studies. In PFC excitatory neurons of living study participants, higher expression of the genes in TPAWN tracked with higher expression of RNA transcripts that in rodent PFC samples are markers of a class of excitatory neurons that connect the PFC to deep brain structures. TPAWN was further reproduced by RNA transcript expression patterns differentiating living PFC samples from postmortem PFC samples, and significant differences between living and postmortem PFC samples were additionally observed with respect to (1) the expression of most primary RNA transcripts, mature RNA transcripts, and proteins, (2) the splicing of most primary RNA transcripts into mature RNA transcripts, (3) the patterns of co-expression between RNA transcripts and proteins, and (4) the effects of some DNA sequence variants on RNA transcript and protein expression. Taken together, this report highlights that studies of brain tissue obtained in a safe and ethical manner from large cohorts of living individuals can help advance understanding of the multiomic foundations of brain function.

## Introduction

An aim of neuroscience is to advance knowledge of how higher-order brain functions in humans originate within tissue from the prefrontal cortex (PFC) and other specialized brain areas. For the purposes of this report, higher-order brain functions are defined as functions that emerge from specialized brain areas such as the PFC through the integration of multiple streams of information that come from other brain areas^1^. Examples of higher-order functions include thought, perception, emotion, and language. Generally speaking, tissue functions originate from deoxyribonucleic acid (DNA), ribonucleic acid (RNA), and protein. Specialized sequences of DNA (“genes”) are transcribed into raw forms of RNA (“primary RNA transcripts”), which are cut and pasted (“spliced”) into processed forms of RNA (“mature RNA transcripts”) that are then translated into proteins^2^. Transcription, splicing, translation, and protein activity are all regulated through an untold number of coordinated processes involving many thousands of DNA sequence variants in the genome, RNA transcripts in the transcriptome, and proteins in the proteome – collectively, these processes are referred to in this report as the “multiomic foundations” of tissue function^3^. In human PFC tissue, multiomic foundations set the stage for neurotransmission (i.e., the electrical transfer of information between neurons), and from complex patterns of neurotransmission involving the PFC and other brain areas emerge higher-order brain functions^4^.

Historically, studies of the brain in living humans during neurosurgical procedures have played an important role in establishing what is currently known about how higher-order brain functions emerge from brain tissue. Studies led by Wilder Penfield, for instance, resulted in the development of the cerebral homunculus that is used in neuroscience education to teach the anatomical basis of higher-order human brain functions^5,6^. Using modern biotechnology, neuroscientists are able to conduct multiomic characterizations of human brain tissue. Yet, the multiomic foundations of neurotransmission in humans, and by extension the multiomic foundations of higher-order human brain functions, remain poorly characterized. This may, in part, be because most of the studies that have used these technologies to characterize human brain tissue have used tissue obtained in the postmortem state^7–12^, which is defined by the cessation of neurotransmission and higher-order brain functions^13^.

For the Living Brain Project, a safe and scalable procedure was developed to acquire PFC tissue from living people for biomedical research purposes (***Figure 1***)^14–17^. Three previous reports on data or samples obtained for the LBP have made the following contributions: (1) using bulk RNA sequencing (bulk RNA-seq; i.e., the capture and quantification of pooled RNA from the cells of a sample), differences in RNA transcript expression between living and postmortem human PFC tissues were characterized and analyses were presented that investigated whether these differences impact the findings of RNA transcript expression studies that only use postmortem tissues^14^; (2) using single-nucleus RNA sequencing (snRNA-seq; i.e., the capture and quantification of RNA from many individual nuclei in a tissue sample), the differences in RNA transcript expression between living and postmortem human PFC tissues were dissected and an approach to statistically account for these differences in analyses of data from postmortem samples was developed^17^; (3) using intracranial measures of neurotransmission obtained while participants played a computer game that engaged higher-order brain functions, patterns of dopamine and serotonin fluctuation in the basal ganglia were identified that associated with social cognition^16^. Collectively, these three prior LBP reports – along with several other recent reports from other research groups^18,19^ – highlight the potential for multiomic studies of brain tissue from living people to help advance understanding of the multiomic foundations of neurotransmission and, by extension, of higher-order brain functions.

**Figure 1.**
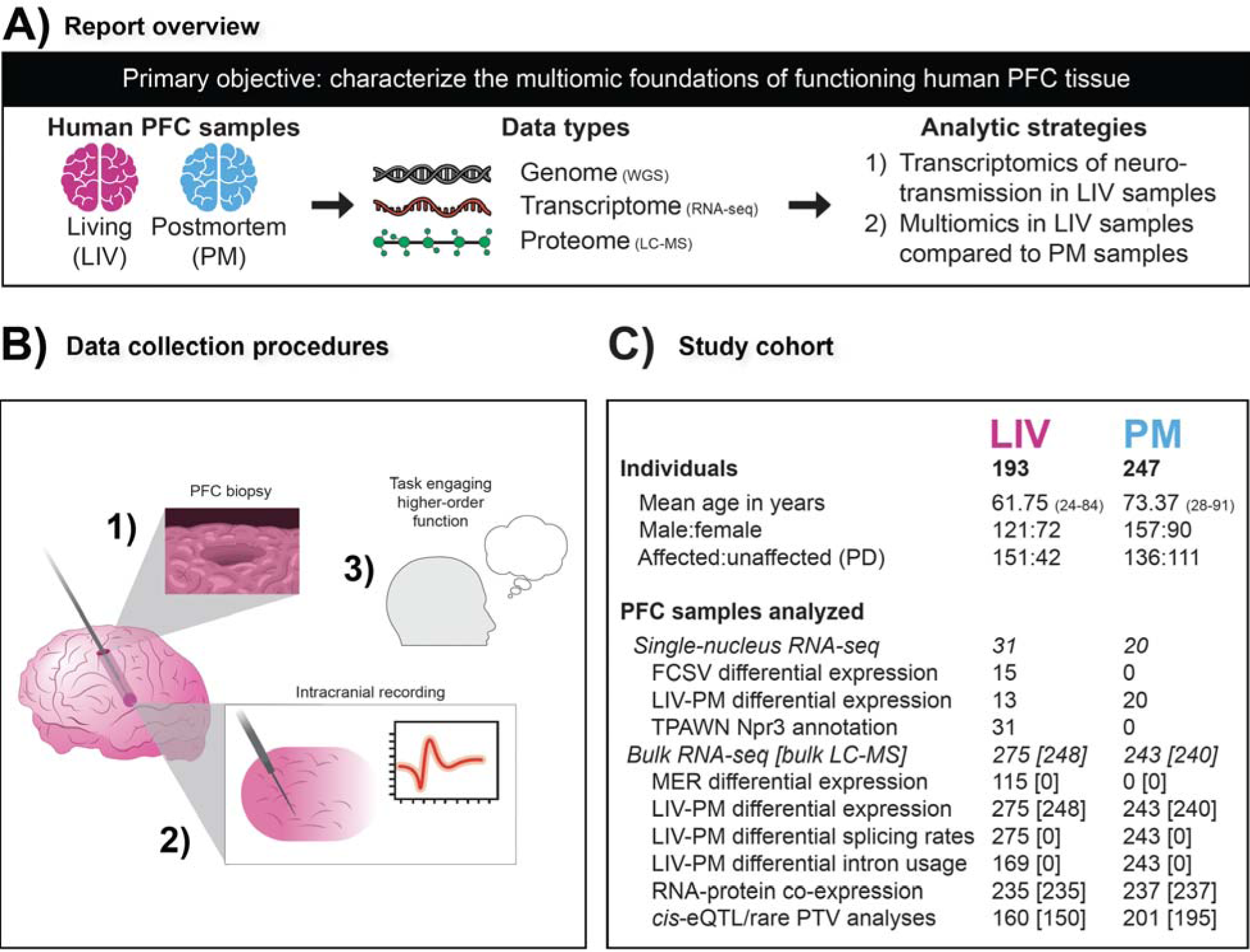
(A) Report overview. Schematic illustrating the study objective and how it was achieved. <u>(B) Data collection procedures.</u> Schematic illustrating how prefrontal cortex (PFC) biopsies are obtained in conjunction with intracranial electrical recordings. In 1) the state of the PFC after the biopsy is obtained is depicted. In 2) a probe obtaining electrical recordings from deep brain structures is depicted. In 3) a cartoon of a person is shown to illustrate that in some cases while electrical recordings are obtained the study participant is engaged in a cognitive task. <u>(C) Study cohort.</u> Numbers refer to sample size (i.e., individuals or samples) except for age. Sample sizes are shown for most of the analyses of LBP data presented in the report. Sample sizes inside and outside of the square brackets indicate counts for bulk RNA-seq and bulk LC-MS analyses, respectively.

Here, towards this end, the following question is addressed using both newly and previously described LBP data: “Does PFC tissue from individuals with intact higher-order brain function have characteristic multiomic foundations?” To address this question, two complementary strategies were employed. The first strategy was to identify in PFC samples obtained from living participants a signature of RNA transcript expression associated with neurotransmission measured intracranially at the time of PFC sampling, in some cases while participants performed a task engaging higher-order brain functions (***Figure 1***). The second strategy was to perform multiomic comparisons between PFC samples obtained from individuals with intact higher-order brain function at the time of sampling (i.e., living study participants) and PFC samples obtained in the postmortem state.

## Results

### Living Brain Project cohort

As first described in the flagship LBP report by Liharska *et al.*^14^, a procedure was developed for the LBP to obtain PFC samples from individuals with intact higher-order brain function at the time of sampling (i.e., PFC samples from living study participants) for research purposes during neurosurgical procedures for deep brain stimulation (DBS), an elective treatment for neurological and mental illnesses^20^. This procedure allows PFC tissue to be sampled in conjunction with other research activities that can take place during DBS surgeries, such as intracranial recordings of neurotransmission and tasks that engage higher-order brain functions (***Figure 1B***). A total of 319 PFC biopsies (“LIV samples”) from 193 living participants were studied for the current report, including unilateral biopsies from 67 participants (49 from the left hemisphere and 18 from the right hemisphere) and bilateral biopsies from 126 participants. For comparison to LIV samples, a cohort of postmortem PFC samples (“PM samples”, N = 247) was assembled from three brain banks. To the extent that it was possible, PM samples were matched to LIV samples for age, sex, and the size of tissue used for RNA and protein extraction. The majority of samples were obtained from individuals with Parkinson’s disease (PD), the most common indication for DBS. All of the LIV samples and PM samples studied in the current report have been studied in previous LBP reports, though the current report introduces several new LBP datasets linked to these samples.

The analyses presented in the current report center around the following eight LBP datasets (***Figure 1C***): (1) single-nucleus RNA sequencing (snRNA-seq) data from 51 PFC samples (31 LIV samples and 20 PM samples) introduced in the LBP report by Vornholt *et al.*^17^; (2) neurotransmission data measured intracranially using fast-scan cyclic voltammetry (FSCV) during 15 DBS surgeries, a subset of which was introduced in the LBP report by Batten *et al.*^16^; (3) bulk RNA-seq data from 518 PFC samples (275 LIV samples and 243 PM samples) introduced in the LBP report by Liharska *et al.*^14^; (4) neurotransmission data measured intracranially using microelectrode recordings in 115 DBS surgeries (this dataset is introduced in the current report); (5) bulk liquid chromatography-mass spectrometry (bulk LC-MS; i.e., the capture and quantification of pooled protein from the cells of a sample) data from 488 PFC samples (248 LIV samples and 240 PM samples; this dataset is introduced in the current report); (6) whole-genome sequencing data from the majority of individuals with PFC samples characterized using bulk RNA-seq and bulk LC-MS (this dataset is introduced in the current report); (7) immunohistochemistry data from 458 PFC samples, a subset of which was introduced in Liharska *et al.*^14^; (8) clinical data introduced in Liharska *et al.*^14^. Each of these eight LBP datasets is more fully described in the LBP report that first introduced the dataset (when applicable), the respective section of the current report that first reports results of analyses of the dataset, and/or in the methods section of the current report. The details regarding the procedures used for data quality control and statistical modeling these data are fully described in the methods section of the current report (***Supplementary Figures 1-4***).

### Differential expression signatures of neurotransmission

#### Section introduction

Higher-order brain functions that emerge from the human PFC ultimately do so from neurotransmission in the PFC and connected brain regions. In living individuals, neurotransmission can be measured directly in these brain regions during neurosurgical procedures using a variety of intracranial electrical recording devices. When such recordings are obtained in conjunction with a brain tissue biopsy, differential expression (DE) can be performed to characterize the association between intracranial electrical recording measurements and the expression levels of individual RNA transcripts (“features”). In DE analysis, for a given set of features (e.g., all RNA transcripts expressed in a given cell type in snRNA-seq data), the expression level of every feature is tested for association with a trait of interest using a regression model. For a given feature, the regression model beta (by convention, the “logFC” value) for the trait of interest captures both the magnitude and direction of the feature-trait association. In this report, the set of feature-trait logFCs for all features tested in a DE analysis is referred to as the “DE signature” of the trait, and features with statistically significant associations with the trait are referred to as “differentially expressed features” (DEFs).

The first strategy to investigate whether the PFC of individuals with intact higher-order brain function has characteristic multiomic foundations was to integrate results from “neurotransmission DE analyses” (i.e., DE analyses where the traits of primary interest are intracranial electrical recording measurements) performed on three independent cohorts: (1) the LBP cohort where FSCV was used to measure intracranial neurotransmission; (2) the LBP cohort where MERs were used to measure intracranial neurotransmission; (3) a cohort from a study published by Berto *et al.*^19^ where intracranial electroencephalography (iEEG) was used to measure intracranial neurotransmission. The DE signatures and DEFs resulting from these three types of neurotransmission DE analyses will be referred to below by the technique used to measure neurotransmission (e.g., “FSCV DE signature,” “FSCV DEFs”). DEFs with positive associations between RNA transcript expression and neurotransmission metrics will be referred to below as “Up DEFs” (e.g., “iEEG Up DEFs”) and DEFs with negative associations between RNA transcript expression and neurotransmission metrics will be referred to below as “Down DEFs” (e.g., “iEEG Down DEFs”).

#### Identification of FSCV DE signatures

FSCV is a method for measuring the fluctuations of dopamine, serotonin, and norepinephrine concentrations in human brain tissue^21–27^. For the current report, FSCV recordings were made during 15 DBS surgeries while study participants (N = 9) played 60 rounds of the ultimatum game^28,29^ approximately 10 minutes after a PFC biopsy was obtained. The FSCV recordings were made from the substantia nigra pars reticulata (SNr), a deep brain structure with direct white matter connections to the PFC in humans^30,31^. In the ultimatum game, the participant must decide whether to accept or reject monetary offers from an avatar (the “proposer”). In each round, the proposer offers a split of $20 to the participant that could range from $1 to $9. If the study participant accepts the offer, they get the offered amount and the proposer gets the remaining amount (i.e., the difference between $20 and the offer). If the participant rejects the offer, neither the participant nor the proposer receives any money. The ultimatum game engages higher-order brain functions such as social reasoning (e.g., determining what constitutes a fair or unfair offer).

From each of the 15 FSCV recordings, a metric was derived for dopamine, serotonin, and norepinephrine signals was by calculating the beta coefficient from a linear model testing the association between fluctuations of the neurotransmitter and the size of the offer made by the proposer to the participant. snRNA-seq was performed on the 15 PFC biopsies obtained in the minutes prior to FSCV recordings, and after the completion of snRNA-seq data quality control procedures 48,735 cells annotated to six brain cell types remained for analysis (19,817 excitatory neurons [Exc]; 4,899 inhibitory neurons [Inh]; 16,062 oligodendrocytes [Oli]; 4,316 astrocytes [Ast]; 2,342 microglia [Micro]; 1,299 oligodendrocyte progenitor cells [OPCs]). For each cell type, DE was performed testing the association between the expression of each RNA transcript detected in that cell type with each of the three neurotransmitters, resulting in a total of 18 FSCV DE signatures. Few FSCV DEFs were identified when using adjusted p-values to define DEFs (the full 18 FSCV DE signatures discovered in this report are provided in ***Supplementary Table 1***). However, using the π_1_ statistic^32^, which provides a lower bound of the proportion of RNA transcripts tested that truly deviate from the null hypothesis of no association between RNA transcript expression and the DE trait of interest (e.g., the FSCV metric), it was estimated that with larger samples sizes FSCV DEFs would be identified (i.e., π_1_ > 0) for 13 of the 18 FSCV DE signatures (***Figure 2A***). Downstream analyses of FSCV DE signatures only utilized “high-confidence” FSCV DE signatures, which were defined as those with a π_1_ statistic greater than 0.10 (***Figure 2A***), and a nominally significant p-value (unadjusted p-value < 0.05) was used to define FSCV DEFs in these signatures. The FSCV DE signature with the strongest signal (i.e., the greatest π_1_ statistic) was the FSCV DE analysis of dopamine measurements and excitatory neuron RNA transcript expression (“dopamine Exc FSCV DE analysis”; π_1_ = 0.41; ***Figure 2B***). Biological pathways enriched in FSCV DEFs were identified using the Kyoto Encyclopedia of Genes and Genomes (KEGG) database, which groups molecular features into sets based on curated annotations from public resources and published literature^33^. KEGG enrichment was performed on the two DEF sets from each of the high-confidence FSCV DE signatures (i.e., Up DEFs and Down DEFs), and the KEGG sets found to be enriched in at least one FSCV DEF set were ‘glutamatergic synapse,’ ‘serotonergic synapse,’ ‘GABAergic synapse,’ ‘cholinergic synapse,’ ‘long-term potentiation,’ ‘long-term depression,’ ‘ion channels,’ ‘neuroactive ligand-receptor interaction,’ ‘circadian entrainment,’ ‘retrograde endocannabinoid signaling,’ ‘oxytocin signaling pathway,’ ‘GnRH secretion,’ ‘chromosome and associated proteins,’ ‘spliceosome,’ ‘exosome’, and ‘protein kinases’ (full KEGG enrichment statistics for the FSCV DE signatures are provided in ***Supplementary Table 2***).

**Figure 2.**
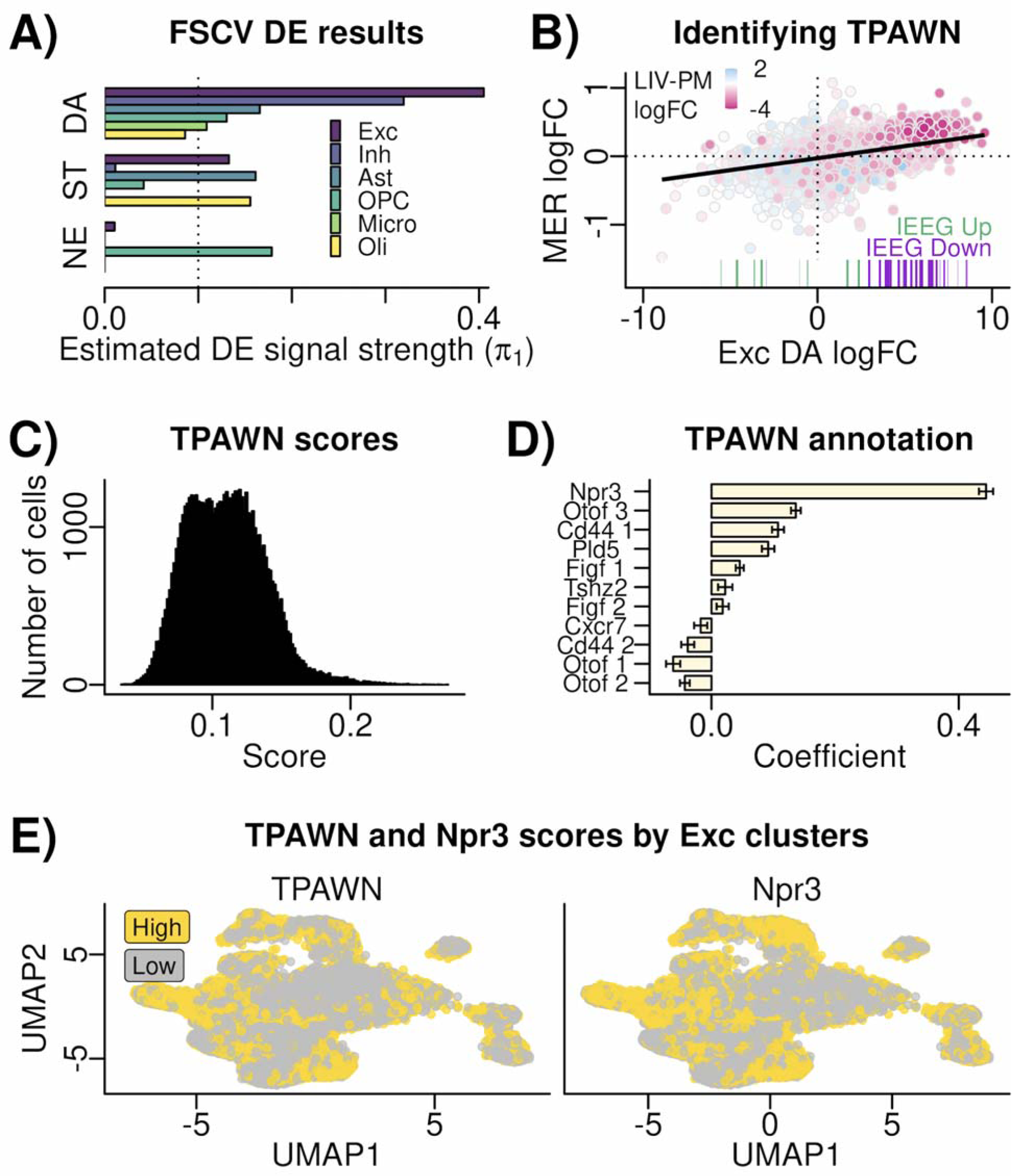
(A) FSCV DE results. Bar plot summarizing the results of the 18 FSCV DE analyses. The result of each DE analyses is summarized by the π_1_ statistic (x-axis), which provides a lower bound of the proportion of RNA transcripts tested that truly deviate from the null hypothesis of no association between RNA transcript expression and the FSCV metric indicated on the y-axis. For each FSCV metric (i.e., each neurotransmitter) there is a result (a bar) shown for DE of that metric performed in each of 6 cell types (represented by different colors) in the snRNA-seq data. <u>(B) Identifying TPAWN.</u> Scatter plot showing relationship of the logFC values from the FSCV DE signature with the greatest signal strength in panel A (x-axis) to the logFC values from the STN MER DE signature (y-axis). The color of points indicates the logFC values from the Exc LIV-PM DE signature. The colors range from pink (greater expression in LIV samples compared to PM samples) to white (no difference in expression between PM samples and LIV samples) to blue (expression greater in PM samples relative to LIV samples). The colored bar beneath select points in the scatter plot indicates whether the RNA transcript represented by the point is an iEEG Up DEF (green bar) or an iEEG Down DEF (purple bar) for the delta wave DE signature in the Berto *et al.* study. <u>(C) TPAWN scores.</u> Histogram showing the distribution of TPAWN scores (x-axis) in 60,084 Exc nuclei from 31 LIV samples with snRNA-seq data. <u>(D) TPAWN</u> <u>annotation.</u> Results of linear regression testing the association between TPAWN scores (dependent variable) and scores reflecting the activity of genes reported by Lui *et al.* to be markers of PFC excitatory neuron projection patterns in mice. The y-axis indicates the different types of PFC projection excitatory neurons in mice using the marker naming scheme in Lui *et al.*. The x-axis is the beta coefficient between the signature indicated on the y-axis and the TPAWN scores from the linear model. Error bars indicate the 95% confidence interval. <u>(E)</u> <u>TPAWN and Npr3 scores by Exc clusters.</u> Scatter plot showing the clustering of 60,084 Exc nuclei from 31 LIV cells. The gene expression data from these cells was reduced to two dimensions using the UMAP approach (shown on the x-axis and y-axis). In the left plot, the points are colored by whether the TPAWN score in the nucleus was high or low. In the right plot, the points are colored by whether the Npr score in the nucleus was high or low. For both plots, the categorization of a nucleus as high or low was done based on whether its score was greater than or less than the mean. FSCV – fast scane cyclic voltammetry; DE – differential expression; DA – dopamine; ST – serotonin; NE – norepinephrine; Exc – excitatory neurons; Inh – inhibitory neurons; Ast – astrocytes; OPC – oligodendrocyte progenitor cells; Micro – microglia; Oli – oligodendrocytes; TPAWN – transcriptional program associated with neurotransmission; MER – microelectrode recording; iEEG – intracranial electroencephalography.

#### Replication of FSCV DE signatures

Given the lack of power in the FSCV DE analyses (i.e., the lack of FSCV DEFs when using adjusted p-values to define FSCV DEFs), it was important to assess using independent data whether the high-confidence FSCV DE signatures truly capture information about the multiomic foundations of neurotransmission in the human brain. Towards this end, neurotransmission DE was performed in two independent datasets (one first introduced in this report, and one previously reported by a different group of researchers).

The first independent dataset was from the subset of the DBS surgeries where bulk RNA-seq had been performed on the PFC biopsy obtained during the surgery and MERs (i.e., a technology for measuring neurotransmission intracranially that is distinct from FSCV) were obtained for analysis along white matter tracts between the PFC biopsy site and the anatomical target for DBS (i.e., subthalamic nucleus [STN] or globus pallidus internus [GPi]) approximately 10 minutes following the PFC biopsy (***Figure 1B***). From these MER data, neurotransmission was represented quantitatively as the aperiodic exponent, which is a metric derived from local field potentials that captures the balance between neuronal inhibition (larger aperiodic exponent values) and excitation (smaller aperiodic exponent values) in the brain tissue surrounding the microelectrode^34^. MERs were analyzed from 115 DBS surgeries (72 surgeries where the STN was the DBS target and 43 surgeries where the GPi was the DBS target) where bulk RNA-seq from the PFC biopsy obtained during the surgery was available for analysis. For each DBS target, DE was performed between the MER metric and mature RNA transcript expression in the PFC (the full MER DE signatures are in ***Supplementary Table 3***). When using adjusted p-values to defined MER DEFs, none were identified, but using the π_1_ statistic^32^ it was estimated that with larger samples sizes MER DEFs would be identified for both the STN (π_1_ = 0.30) and the GPi (π_1_ = 0.29) MER DE signatures. Comparing each of these two MER DE signatures to each of the 10 high-confidence FSCV DE signatures using Spearman’s correlation revealed a significant correlation after multiple test correction for 80% of the comparisons (i.e., 16 of the 20 comparisons between one of the 10 FSCV DE signatures and one of the two MER DE signatures). The strongest correlation was observed between the dopamine Exc FSCV DE signature and the STN MER DE signature (Spearman’s ρ = 0.30; adjusted p-value = 2.05 x 10^−33^; ***Figure 2B***).

The second independent dataset analyzed to assess whether the FSCV DE signatures truly capture information about the multiomic foundations of neurotransmission in the human brain was from the previously published study by Berto *et al.* of 16 individuals undergoing temporal cortex resection to treat epilepsy^19^. The Berto *et al.* study was of the following design: (1) iEEG recordings (i.e., a technology for measuring neurotransmission intracranially that is distinct from both FSCV and MER) were made of the temporal cortex while participants successfully encoded memories, and from each iEEG recording a metric was derived summarizing each of six frequencies of neural oscillations (alpha, beta, delta, gamma, high gamma, and theta); (2) days after the iEEG recordings, temporal cortex tissue was resected for therapeutic purposes during a neurosurgical procedure; (3) RNA sequencing of the resected temporal cortex tissue was performed; (4) DE was performed for each of the six frequencies of neural oscillations derived from the iEEG recordings. The six lists of iEEG DEFs reported in this published study were downloaded, and each list was split into two lists – one for iEEF Up DEFs and one for iEEF Down DEFs. For each of the 10 high-confidence FSCV DE signatures analyzed, the FSCV Up DEFs and the FSCV Down DEFs were tested for enrichment of the 12 iEEG DEF lists (for a total of 240 tests). After accounting for multiple tests, 6 comparisons survived correction. The most significant enrichment was between the dopamine Exc FSCV Up DEFs and the delta wave iEEG Down DEFs (Fisher’s test OR = 6.30, adjusted p-value = 4.56 x 10^−45^). A significant enrichment was also seen for dopamine Exc FSCV Down DEFs and the delta wave iEEG Up DEFs (Fisher’s test OR = 3.50, adjusted p-value = 2.56 x 10^−15^) (***Figure 2B***).

#### Defining a transcriptional program associated with neurotransmission (TPAWN)

Given the different designs of the studies that identified the FSCV, MER, and iEEG DE signatures, the findings presented in this section thus far suggest a shared set of RNA transcripts may be involved in regulating neurotransmission in the human brain in different contexts. To test this hypothesis, a set of RNA transcripts was identified that had evidence for association to neurotransmission from multiple types of neurotransmission DE analyses, and this set of transcripts was characterized using multiple types of external data sources.

To identify the set of transcripts, the following procedure was performed, with input to the procedure being DEFs from the 10 high-confidence FSCV DE signatures, the two MER DE signatures, and the six iEEG DE signatures: (1) the DEFs for every DE signature were compared to the DEFs of every other DE signature using Fisher’s exact tests; (2) two DE signatures were defined as “connected” to one another if a Fisher’s exact test comparing the DEFs of the two signatures was significant after multiple test correction; (3) a network was constructed where nodes were DE signatures and edges were defined by the previous step; (4) a fully connected sub-component of the network containing at least one DE signature from each of the three types of neurotransmission DE analyses (i.e., FSCV, MER, and iEEG) was identified; (5) RNA transcripts that were DEFs in DE signatures from at least two of the three types of neurotransmission DE analyses in the fully connected subcomponent were identified. This procedure output a set of 588 RNA transcripts suspected to be involved in what will be referred to in the remainder of this report as a “transcriptional program associated with neurotransmission” (TPAWN; ***Supplementary Table 4***).

To support the notion that TPAWN genes (i.e. the genes encoding the RNA transcripts in TPAWN) play a role in neurotransmission and higher-order human brain function, four analyses were performed. First, the evolutionary constraint of TPAWN genes was examined, as more constrained genes have been shown to be enriched for genes implicated in disorders of higher-order brain function^35^. Relative to all genes expressed in excitatory neurons, TPAWN genes were evolutionarily constrained as measured by having lower “loss-of-function observed/expected upper bound fraction” (LOEUF) scores (K-S test p-value = 1.87 x 10^−9^). Second, TPAWN genes were compared to genes implicated in neurotransmission through experiments of model systems, and found to be enriched with genes associated to neurotransmission in cell lines or animal models by (1) the NeuroExpresso/NeuroElectro study^36^ (OR = 2.88, p-value = 1.53 x 10^−9^) and (2) a report on data from the Allen Institute for Brain Science^37^ (OR = 1.80, p-value = 2.21 x 10^−12^). Third, genes implicated in human health and disease through common genetic variation in the GWAS catalog were tested for enrichment of TPAWN genes, and after multiple test correction TPAWN genes were found to enriched for genes linked to brain-related disorders (e.g., schizophrenia, bipolar disorder), brain-related traits (e.g., cognition, educational attainment), behaviors (e.g., risk-taking), measures of brain tissue pathology (e.g., plaques), and brain imaging phenotypes (e.g., cortical thickness) (***Supplementary Table 5***). Fourth, whole-exome sequencing data from 29,064 ancestrally diverse individuals in a general health system population in New York City was mined to investigate the consequences of rare protein-truncating variants (PTVs) in genes encoding RNA transcripts in TPAWN on higher-order brain functions. This was done by performing a mental health-specific phenome-wide association test on rare PTV carrier status. After multiple test correction for the number of mental health phecodes examined, a significant increase in risk for ‘hallucinations’ (phecode 292.6) was observed in carriers of rare PTVs in TPAWN genes compared to non-carriers (OR = 3.81, unadjusted p-value = 5.62 x 10^−4^, adjusted p-value = 0.04).

The FSCV and MER DE analyses that led to the identification of TPAWN both involved RNA-seq of PFC samples and intracranial recordings of deep brain structures, suggesting that TPAWN may capture molecular processes occurring within PFC excitatory neurons that communicate molecular information to deep brain structures through axonal projections. To test this hypotheses, a series of analyses were performed on the Exc snRNA-seq data from LIV samples (N = 31 sample; includes the 15 with FSCV data). For each Exc nucleus, a score was created that captured the activity of TPAWN genes in that nucleus (where higher scores reflected higher expression levels for TPAWN genes). Visualizing the resulting scores, a bimodal distribution was observed (***Figure 2C***). In a recent study, Lui *et al*. characterized seven subtypes of PFC excitatory neurons in rodents and annotated these subtypes with respect to their axonal long-range projections throughout the brain^38^. A score for each of the 7 subtypes was calculated for the Exc cells in the LBP snRNA-seq data, and the association between the TPAWN scores and these 7 subtype scores was assessed using a linear model that included all 7 subtype scores as independent variables. This analysis found the subtype labeled Npr3 in Lui *et al.* had the strongest association to TPAWN scores, with the regression model coefficient for this variable over 3.5-fold the magnitude of the regression model coefficient for the next strongest associated variable (***Figure 2D***). After accounting for any overlap between TPAWN genes and Npr3 markers, Exc with high TPAWN and Npr3 scores were not isolated in a single subcluster of Exc cells (***Figure 2E***). Lui et al determined that Npr3 cells were preferentially located in layer 5 of the PFC and had projections to many of the deep brain structures examined in their study. Based on these observations, TPAWN in the PFC identified with FSCV data from the SNr may be driven by subcortical-projecting Exc cells.

### Multiomic comparisons between LIV samples and PM samples

#### Section introduction

As stated at the outset of this report, the objective of this report is to examine whether PFC tissue from individuals with intact higher-order brain function has characteristic multiomic foundations, and two strategies were utilized towards this end. So far, the results have been presented from the first strategy, which was to identify in PFC samples obtained from living participants a signature of RNA transcript expression associated with neurotransmission measured intracranially at the time of PFC sampling (***Figure 1***). The second strategy was to perform multiomic comparisons between PFC samples obtained from individuals with intact higher-order brain function at the time of sampling (i.e., LIV samples) and PFC samples obtained in the postmortem state (i.e., PM samples) (***Figure 1C***).

#### Neurotransmission DE signatures are reproduced in LIV-PM DE signatures

An assumption inherent to using analyses comparing LIV samples to PM samples to accomplish the overall objective of the report is that differences between LIV samples and PM samples capture the multiomic foundations of neurotransmission and, by extension, of higher-order brain functions. To directly test this assumption, DE of “LIV-PM status” (i.e., whether a PFC sample is a LIV sample or a PM sample) was performed for each cell type in snRNA-seq data from 13 LIV samples and 20 PM samples and the concordance between the 10 high-confidence FSCV DE signatures and the resulting LIV-PM DE signatures was assessed. The 13 LIV samples in these LIV-PM DE analyses were separate from the 15 LIV samples in the FSCV DE analyses. After multiple test correction, for 9 of the 10 FSCV DE signatures a significant correlation was observed between the FSCV DE signature and the snRNA-seq LIV-PM DE signature in the corresponding cell type, and for 8 of these 9 FSCV DE signatures the direction of the correlation between the FSCV DE signature and the corresponding LIV-PM DE signature showed that the features associated with greater FSCV metrics were overlapping with features more highly expressed in LIV samples compared to PM samples. The strongest correlations were the dopamine FSCV DE signatures in PFC neurons (Spearman’s ρ = −0.44 for Inh and −0.42 for Exc; adjusted p-values < 2.2 x 10^−16^; ***Figure 2B***). These observations suggest that molecular differences between LIV samples and PM samples capture the multiomic foundations of neurotransmission and establish rationale for the remaining analyses presented in this report.

#### Identifying RNA transcript and protein expression levels characteristic of LIV samples

Utilizing the bulk RNA-seq data from (N = 518 PFC samples [275 LIV samples and 243 PM samples]) and bulk LC-MS data (N = 488 PFC sample [248 LIV samples and 240 PM samples]; 472 of which were in the 518 samples with bulk RNA-seq data), expression levels were quantified for 22,955 primary RNA transcripts, 30,099 mature RNA transcripts, and 6,415 proteins (***Figure 3A***). LIV-PM DE was performed separately for primary RNA transcripts, mature RNA transcripts, and proteins, and the three resulting LIV-PM DE signatures with corresponding KEGG enrichment test results are provided in ***Supplementary Tables 6 and 7***. By convention for this report for LIV-PM DE signatures, positive logFC values represent higher expression in PM samples compared to LIV samples and negative logFC values represent higher expression in LIV samples compared to PM samples. LIV-PM DEFs (i.e., DEFs identified in LIV-PM DE analyses) with negative logFC values are referred to as LIV DEFs and LIV-PM DEFs with positive logFC values are referred to as PM DEFs. Significant differences in expression were observed between LIV samples and PM samples for 74.03% of primary RNA transcripts (8,892 LIV DEFs and 8,102 PM DEFs), 70.20% of mature RNA transcripts (13,313 LIV DEFs and 7,817 PM DEFs), and 60.81% of proteins (1,898 LIV DEFs and 2,003 PM DEFs) (***Figure 3A, Row 3***). A significant Spearman’s correlation coefficient was observed when comparing the LIV-PM DE signatures from (1) primary and mature RNA transcripts (ρ = 0.45, p-value < 2 x 10^−16^) and (2) mature RNA transcripts and proteins (ρ = 0.18, p-value < 2 x 10^−16^) (***Figure 3B***). To establish that the LIV-PM DE signatures identified in this section are not explained by variables with the potential to confound measures of RNA transcript or protein expression, analyses were performed assessing the stability of the LIV-PM DE signatures with respect to the following 12 potential confounding variables: (1) data generation batch; (2) institution of origin of the PM samples; (3) postmortem interval (PMI); (4) diagnosis of PD in living participants and postmortem donors; (5) severity of PD symptoms in living participants; (6) dose of dopamine replacement therapy in living participants; (7) neuropathology in LIV and PM samples; (8) type and dose of anesthesia administered to living participants during DBS surgery; (9) method of LIV sample preservation upon collection during DBS surgery; (10) RNA integrity number (RIN); (11) age differences between living participants and postmortem donors; (12) cell type composition differences between LIV samples and PM samples. The LIV-PM DE signatures were stable with respect to all 12 potential confounding variables, and an extended presentation of these analyses are presented in the ***Supplementary Information***.

**Figure 3.**
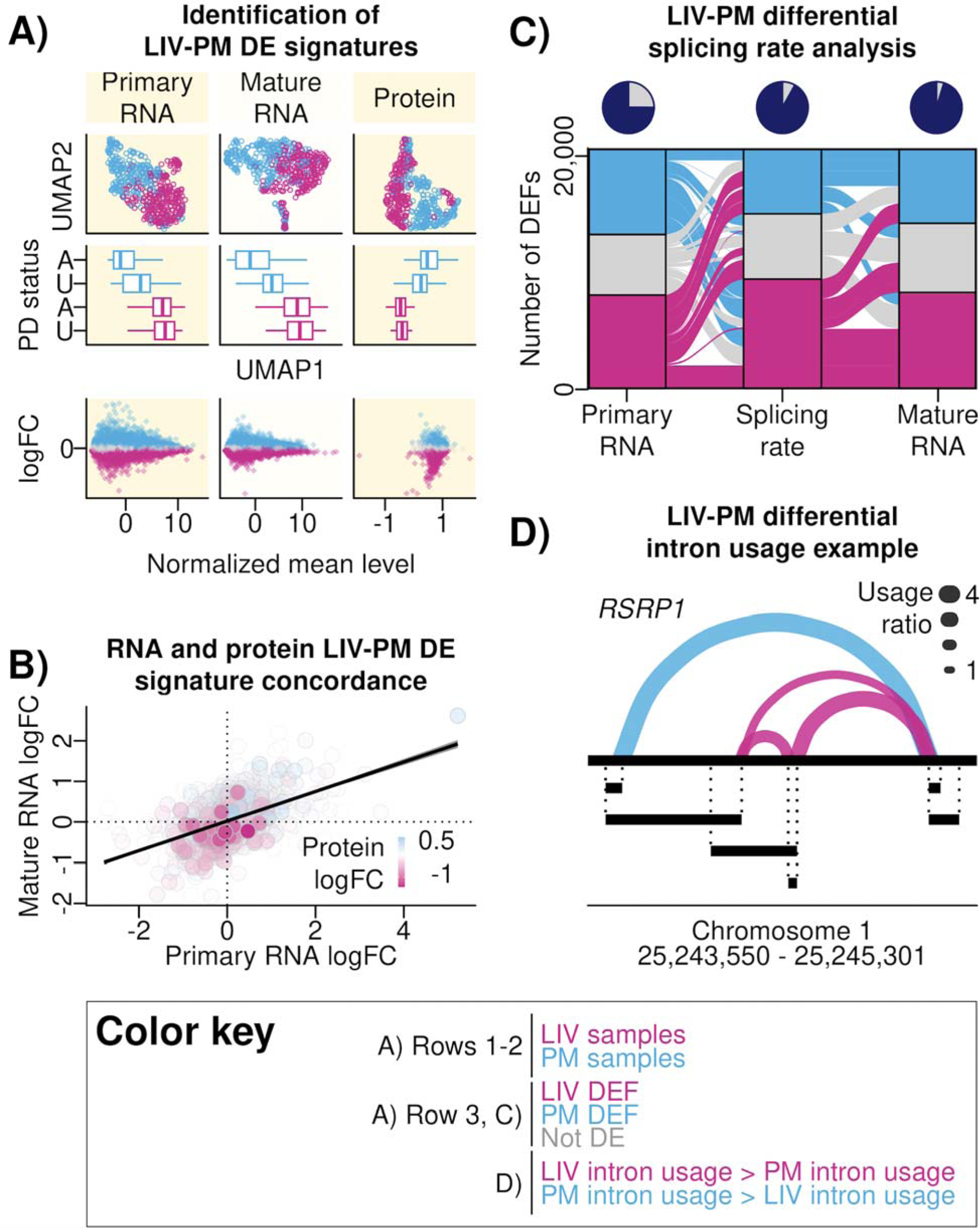
(A) Identification of LIV-PM DE signatures. A matrix of nine plots is presented that altogether summarize the differences in RNA and protein expression levels between LIV samples and PM samples. Each row of the matrix is a different analysis, and each column is a different data type (primary RNA, mature RNA, protein). The off-white background is to differentiate columns from one another. <u>Top Row:</u> scatter plots showing the results of dimensionality reduction performed on primary RNA, mature RNA, and protein expression data using the UMAP algorithm. Each point is a sample, and colors differentiate LIV samples (pink) from PM samples (blue). The horizontal axis is the first UMAP dimension (“UMAP 1”), the vertical axis is the second UMAP dimension (“UMAP 2”). <u>Middle Row:</u> boxplots showing distributions of UMAP 1 values (horizontal axis) stratified on the vertical axis both by LIV-PM status and Parkinson’s disease (PD) status of the samples (LIV samples - pink, PM samples - blue; samples from individuals with PD - A [“Affected”], samples from individuals without PD - U [“Unaffected”]). For each boxplot: the colored line inside the box is the media; the left and right edges of the box are the first and third quartiles (the 25th and 75th percentiles); the right whisker extends from the right edge of the box to the largest y-axis value no further than 1.5 times the interquartile range away from the right edge of the box; the left whisker extends from the left edge of the box to the smallest y-axis value no further than 1.5 times the interquartile range away from the left edge of the box. <u>Bottom Row:</u> scatter plots showing the results of differential expression analysis comparing LIV samples to PM samples for each primary RNA transcript, mature RNA transcript, and protein feature quantified for the bulk RNA-seq and bulk LC-MS analyses in the current report. Each point is a feauture (i.e., RNA transcript or protein). The horizontal axis shows the average normalized RNA transcript or protein expression level. The vertical axis shows the logFC values, and the range of values is −4.95 to 5.29 for primary RNA transcripts, −7.71 to 5.40 for mature RNA transcripts, and −1.71 to 1.27 for protein. Positive logFC values are indicative of higher levels in PM samples compared to LIV samples, and negative logFC values are indicative of higher levels in LIV samples compared to PM samples. Colors differentiate features with levels that were significantly higher in PM compared to LIV (blue), significantly higher in LIV compared to PM (pink), or not significantly different between LIV and PM (gray). <u>(B) RNA and protein LIV-PM DE signature concordance.</u> Scatter plot showing the concordance of RNA transcript and protein LIV-PM DE signatures. Each point is a feature, and only features present in all 3 LIV-PM DE signatures (i.e., primary RNA, mature RNA, protein) are plotted. The x-axis shows the logFC values from the primary RNA transcript LIV-PM DE analysis. The y-axis shows the logFC values from the mature RNA transcript LIV-PM DE analysis. The color of points indicates the logFC values from the protein LIV-PM DE signature. The colors range from pink (greater expression in LIV samples compared to PM samples) to white (no difference in expression between PM samples and LIV samples) to blue (expression greater in PM samples relative to LIV samples). <u>(C) LIV-PM differential splicing</u> <u>rate analysis.</u> An alluvial plot showing changes in the RNA transcript LIV-PM DE signature across three variables (represented by three “pillars” on the horizontal axis) that were tested for differences between LIV and PM samples on the same set of features: levels of primary RNA transcript expression (left pillar), the rate of conversion of primary RNA to mature RNA (middle pillar), and levels of mature RNA expression (right pillar). There are three categories, called “stratum”, within each of the three pillars: RNA features tested that were greater in LIV samples compared to PM samples (pink), greater in PM samples compared to LIV samples (blue), or not significantly different between LIV samples and PM samples (gray). Each “alluvial fan” (i.e., a set of thick wavy lines connecting pillars pillars) shows change of a set of features across the three variables and is comprised of two “flows”, which are the segments connecting adjacent pillars. Flows are colored by the DE status of the stratum from which it originated, and fans can therefore be comprised of flows of multiple colors. The pie charts above each pillar represent the cumulative proportion of the transcriptome that is different between LIV samples and PM samples in at least one of the three levels tested (totaling over 95% as indicated by the proportion of the pie that is colored purple in the last pie chart). <u>(D) LIV-PM differential intron usage</u> <u>example.</u> Differential intron usage between LIV samples and PM samples is shown for the *RSRP1* intron cluster. Exons are represented by the black segments beneath the continuous black line (which represents the full gene). Introns in the cluster are represented by curves that connect two exon ends. The thickness of each curve represents the intron usage ratio. To calculate the intron usage ratio, the mean intron usage was calculated separately for LIV and PM samples and the larger mean intron usage was divided by the smaller mean intron usage. Curves colored pink are introns with greater usage in LIV compared to PM and curves colored blue are introns with greater usage in PM compared to LIV.

#### Identifying RNA splicing and RNA-protein co-expression characteristic of LIV samples

The multiomic foundations of tissue functions are comprised not simply of the expression levels of RNA transcripts and proteins but also by RNA splicing (i.e., the processing of primary RNA into mature RNA) and by RNA-protein co-expression (i.e., the relative expression levels of RNA transcripts to proteins). To help establish these components of the multiomic foundations of PFC function, RNA splicing and RNA-protein co-expression were compared between LIV samples and PM samples using the bulk RNA-seq and bulk LC-MS data. The results of the RNA splicing analyses (i.e., differential splicing rate analysis and differential intron usage analysis) are presented in this section, and the results of the RNA-protein co-expression analyses are presented in the ***Supplementary Information*** (as well as in ***Supplementary Figure 5*** and ***Supplementary Table 8***).

For a given RNA transcript at a single moment in time, the splicing rate (i.e., the amount of mature RNA transcripts relative to the amount of primary RNA transcripts) is an emergent property of transcriptional regulation^39^. Differential splicing rate analysis (i.e., comparing the splicing rate in LIV samples to the splicing rate in PM samples for each RNA transcript expressed) was performed using a regression model that tested the association between LIV-PM status and splicing rates for the 20,671 RNA transcripts with both primary RNA and mature RNA transcript expression detected. Significantly different splicing rates were found for 73.00% of the RNA transcripts tested (9,448 with greater splicing rates in LIV samples compared to PM samples and 5,641 with higher splicing rates in PM samples compared to LIV samples). These included nearly all of the LIV-PM DEFs that had opposite directions of effect in the primary and mature RNA transcript LIV-PM DE signatures (e.g., RNA transcripts that were LIV DEFs in the primary RNA and PM DEFs in the mature RNA; ***Figure 3C***). For 1,074 RNA transcripts, splicing rates were higher in PM samples compared to LIV samples even though the primary and mature RNA transcript expression levels were higher in LIV samples compared to PM samples (i.e., the RNA transcript was a LIV DEF in both the primary and mature RNA transcript LIV-PM DE signatures). Similarly, for 1,635 RNA transcripts, splicing rates were higher in LIV samples compared to PM samples even though the primary and mature RNA transcript expression levels were higher in PM samples compared to LIV samples (i.e., the RNA transcript was a PM DEF in both the primary and mature RNA transcript LIV-PM DE signatures). Altogether, 95.27% of the 20,671 RNA transcripts with both primary RNA and mature RNA transcript expression detected significantly differed between LIV samples and PM samples with respect to either primary RNA transcript expression levels, mature RNA transcript expression levels, or splicing rates (***Figure 3C***).

For an intron in a RNA transcript, the intron usage level is the percentage of mature RNA transcripts that result from splicing the intron out of primary RNA transcripts. For a primary RNA transcript containing multiple introns, using different combinations of introns results in different forms of mature RNA transcripts (“isoforms”), which are translated into different forms of the same protein^40^. For a set of introns in a primary RNA transcript (i.e., an intron cluster), differential intron usage analysis tests whether two groups of samples (e.g., LIV samples and PM samples) differ with respect to patterns of intron usage (and, therefore, patterns of mature RNA transcript isoform abundance)^41^. Intron usage levels were quantified for LIV samples and PM samples from the bulk RNA-seq data (for these analyses, a single LIV sample was retained per living participant for the data to be compatible with the software used; ***Figure 1C***). After data processing and quality control procedures had been completed, 11,222 intron clusters (covering 28,001 introns and mapping to 6,797 unique RNA transcripts) were tested for differential intron usage between LIV samples and PM samples. Significant differences in intron usage were detected for 63.63% of the intron clusters tested (7,141 intron clusters), which covered 65.81% of the introns tested (18,428 introns; 9,649 introns with higher usage rates in LIV samples and 8,779 introns with higher usage rate in PM samples) and 74.15% of the RNA transcripts tested (4,972 RNA transcripts; 97.47% of which significantly differed between LIV samples and PM samples in either the primary RNA transcript, mature RNA transcript, or splicing rate LIV-PM DE signatures). Differential intron usage was most significant for *RSRP1* (adjusted p-value = 5.91 x 10^−162^; ***Figure 3D***), which encodes a component of the spliceosome^42^.

#### Effects of DNA sequence variants on RNA transcript and protein expression in LIV samples

DNA sequence variants (i.e., sites in the human genome where genotypes vary in the population) contribute to higher-order human brain functions and are believed to do so by regulating the expression of RNA transcripts and proteins^43^. To date, most studies of RNA transcript and protein expression regulation by DNA sequence variants in the human brain have used tissues from individuals in the postmortem state^44,45^. In light of the large number of RNA transcripts and proteins differentially expressed between LIV samples and PM samples, analyses were performed to determine if RNA transcript and protein expression regulation by DNA sequence variants differs between LIV samples and PM samples. In the main text of this section, results are presented for analyses of common DNA sequence variants, while in the ***Supplementary Information*** (and ***Supplementary Figure 6***) results are presented for analyses of rare DNA sequence variants.

Common DNA sequence variants are sites in the human genome where the minor allele frequency in the population is greater than an arbitrarily selected threshold (e.g., 1%). In studies that examine molecular feature expression regulation by common DNA sequence variants, for a given molecular feature (i.e., RNA transcript or protein), a test of association is performed between the feature expression level and the common DNA sequence variants located near the transcription start site of the gene that encodes the feature^46^. When a statistically significant association is found, the common DNA sequence variant can be referred to as a “*cis*-expression quantitative trait locus” (*cis*-eQTL) and the feature as a “*cis*-eQTL-regulated feature” (CRF). For the current report, *cis*-eQTLs and CRFs were identified for PFC RNA transcript expression in 361 individuals with WGS and bulk RNA-seq data (161 LIV samples and 200 PM samples) and for PFC protein expression in the 345 individuals with WGS and bulk LC-MS data (150 LIV samples and 195 PM samples). The RNA transcripts and proteins included in these analyses were those with at least one common DNA sequence variant within one mega-base of the transcription start site of the gene encoding the RNA transcript or protein (21,047 primary RNA transcripts, 27,619 mature RNA transcripts, and 5,232 proteins). After multiple test correction, a *cis*-eQTL was identified for 30.71% of primary RNA transcripts (6,463 CRFs), 18.00% of mature RNA transcripts (4,969 CRFs), and 10.76% of proteins (563 CRFs; ***Supplementary Table 9***). CRFs were significantly enriched for CRFs identified in a previous study of postmortem human brain tissue^11^ using Fisher’s exact tests (OR for primary RNA CRFs = 3.67; OR for mature RNA CRFs = 7.92; OR for protein CRFs = 35.18, all p-values < 2.2 x 10^−16^).

Amongst the results presented thus far are two seemingly contradictory observations: (1) the majority of molecular features are differentially expressed between LIV samples and PM samples and (2) the set of molecular features with expression regulated by *cis*-eQTLs identified in the current study is overlapping with sets found in studies that used only postmortem brain samples. These observations suggest the molecular features that remain at stable expression levels regardless of LIV-PM status (i.e., are not LIV-PM DEFs) are enriched for the features most strongly regulated by *cis*-eQTLs. To test this possibility, for each molecular feature, the variances in feature expression explained by (1) common DNA sequence variants and (2) LIV-PM status were calculated independently of one another (allowing them to capture the same variance) and compared. The variances in CRF expression explained by *cis*-eQTLs took on a large range of values, with the expression of some CRFs being controlled almost entirely by *cis*-eQTLs and the expression of other CRFs being controlled more modestly (***Figure 4A***). To determine if LIV-PM DEFs are depleted of the CRFs most strongly regulated by *cis*-eQTLs, an iterative procedure was performed where each iteration calculated the overlap between LIV-PM DEFs and CRFs using a Fisher’s exact test. Successive iterations used increasingly stringent criteria to define CRFs, and definitions were made more stringent with each iteration by increasing the minimum variance in feature expression required to be explained by a *cis*-eQTL for the feature to be considered a CRF. For primary RNA transcripts, mature RNA transcripts, and proteins, as the threshold for defining CRFs became more stringent, CRFs became more depleted of LIV-PM DEFs (***Figure 4A***).

**Figure 4.**
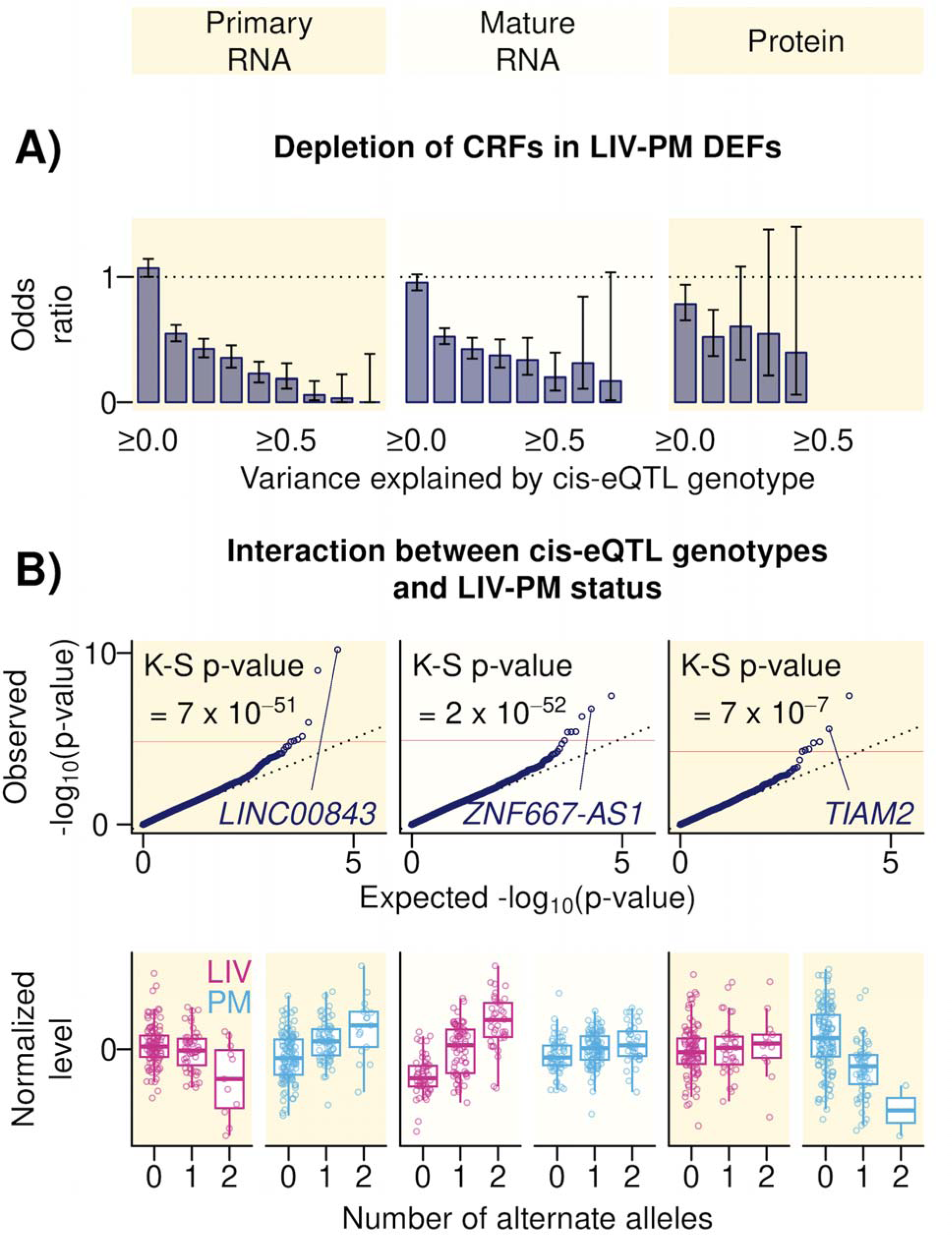
(A) Depletion of CRFs in LIV-PM DEFs. Bar plots showing the results of testing th overlap of LIV-PM DEFs CRFs. To determine if LIV-PM DEFs are depleted of the CRFs most strongly regulated by *cis*-eQTLs, an iterative procedure was performed where each iteration calculated the overlap between LIV-PM DEFs CRFs using a Fisher’s exact test. Successive iterations used increasingly stringent criteria to define CRFs, and definitions were made more stringent with each iteration by increasing the minimum variance in RNA transcript or protein expression required to be explained by a *cis*-eQTL for the RNA transcript or protein to be considered a CRF. The horizontal axis of each bar plot shows the successive iteration. The summary statistic of this test is the odds ratio, presented on the vertical axis of each bar plot. Error bars are the 95% confidence interval of the odds ratio from Fisher’s exact test, and error bars that do not cross the dotted horizontal line intersecting the vertical axis at 1 indicate statistical significance. <u>(B) Interaction between *cis*-eQTL genotypes and LIV-PM status.</u> *Top Row* – Quantile-quantile (QQ) plots of the results of “context-dependent” *cis*-eQTL analyses, where for each primary RNA transcript, mature RNA transcript, or protein a regression model was employed testing for a significant interaction between LIV-PM status and nearby common DNA sequence variant genotypes. Each point represents one RNA transcript or protein. The horizontal axis is the expected distribution of −log_10_(p-values) under the assumption of uniformity and the vertical axis is the observed distribution of −log_10_(p-values). The dotted black line shows the diagonal for reference purposes. The solid red line shows the threshold above which tests were considered statistically significant after multiple test correction. The K-S test p-value shows the statistical significance of the deviation of the observed p-value distribution from the expected p-value distribution. The labeled point in each plot is the gene symbol for the result that is presented in more detail in the bottom row. *Bottom Row* – Boxplots showing for one Primary RNA CD-CRF, one Mature RNA CD-CRF, and one protein CD-CRF the normalized expression levels (vertical axes) in LIV samples (pink points and boxplots) and PM samples (blue points and boxplots) stratified by the number of alternate alleles (horizontal axes) at the CD-cis-eQTL site. There is one boxplot overlaying each set of points. The colored line in each box is the median. The top and bottom edges of each box are the first and third quartiles (the 25th and 75th percentiles). The upper whisker of each box extends from the top edge of the box to the largest y-axis no further than 1.5 times the interquartile range away from the top edge of the box. The lower whisker extends from the bottom edge of the box to the smallest y-axis value no further than 1.5 times the interquartile range away from the bottom edge of the box.

The observation that the sets of CRFs identified in the current study using both living and postmortem brain samples are overlapping with the sets found in studies that used only postmortem brain samples did not rule out the possibility that for a molecular feature the regulatory effect of a common DNA sequence variant may be “context-dependent” (i.e., different based on the LIV-PM status of the samples analyzed). To investigate this possibility, for each molecular feature, a regression model was employed testing for a significant interaction between LIV-PM status and the genotypes at nearby common DNA sequence variants. When a statistically significant association was found, the common DNA sequence variant was referred to as a “context-dependent *cis*-eQTL” (CD-*cis*-eQTL) and the feature as a “context-dependent-CRF” (CD-CRF). After multiple test correction, a total of 21 CD-CRFs were identified (7 each in the primary RNA, mature RNA, and protein analyses; full results of the context-dependent *cis*-eQTL analysis are provided in ***Supplementary Table 11***). Previous studies suggest that context-dependent *cis*-eQTL analyses are underpowered compared to standard *cis*-eQTL analyses performed in studies of similarly sized cohorts^47,48^. Therefore, to assess whether there is evidence the regulatory effects of *cis*-eQTLs on feature expression are dependent on the LIV-PM status of the samples analyzed, the K-S test was performed on the unadjusted p-value distributions of the context-dependent *cis*-eQTL results, and for primary RNA transcript, mature RNA transcript, and protein expression, the unadjusted p-value distributions significantly deviated from uniformity (K-S test p-values: primary RNA = 1.06 x 10^−34^, mature RNA = 6.32 x 10^−158^, protein = 1.52 x 10^−14^; ***Figure 4B, Top Row***). An example of one primary RNA CD-CRF, one mature RNA CD-CRF, and one protein CD-CRF are presented in ***Figure 4B*** (***Bottom Row***).

Given the relationship reported above between the FSCV DE signatures and LIV-PM DE signatures, the depletion of CRFs for LIV-PM DEFs led to the hypothesis that, compared to the expression of other RNA transcripts, the expression of RNA transcripts associated with neurotransmission may be less regulated by common DNA sequence variants. To test this hypothesis, an iterative procedure was performed where each iteration used a Fisher’s exact test to calculate the overlap between mature RNA transcript CRFs and RNA transcripts associated with neurotransmission through either (1) TPAWN, (2) the NeuroExpresso/NeuroElectro study^36^, or (3) a report on data from the Allen Institute for Brain Science^37^. Successive iterations used increasingly stringent criteria to define CRFs, and definitions were made more stringent with each iteration by increasing the minimum variance in mature RNA transcript expression required to be explained by a *cis*-eQTL for the mature RNA transcript to be considered a CRF. As the threshold for defining CRFs became more stringent, CRFs became more depleted of RNA transcripts in TPAWN (***Supplementary Table 10***), and the most significant depletion was observed using a variance explained threshold of 0.1 (Fisher’s exact test OR = 0.08, adjusted p-value = 1.58 x 10^−7^). These observations were reproduced for RNA transcripts associated with neurotransmission by the NeuroExpresso/NeuroElectro study (Fisher’s exact test OR = 0.43, adjusted p-value = 0.01) and for RNA transcripts associated with neurotransmission by a report on data from the Allen Institute for Brain Science (Fisher’s exact test OR = 0.41, adjusted p-value = 3.01 x 10^−28^) (***Supplementary Table 10***).

## Discussion

The main objective of the LBP study reported here was to characterize the multiomic foundations of PFC tissue obtained from individuals with intact higher-order brain function at the time of tissue sampling. To achieve this objective, two complementary strategies were employed. First, RNA transcript expression in PFC samples obtained from individuals with intact higher-order brain function at the time of sampling was tested for association with neurotransmission measured intracranially at the time of PFC sampling, in some cases while individuals performed a task that engaged higher-order brain function. Second, multiomic comparisons were performed between PFC samples obtained from individuals with intact higher-order brain function at the time of sampling and PFC samples obtained from individuals in the postmortem state at the time of sampling. The brain expression levels of a set of approximately 600 RNA transcripts (i.e., TPAWN) was found to associate with intracranial measurements of neurotransmission. These RNA transcripts were (1) enriched for RNA transcripts associated with measures of neurotransmission in rodent and cell models, (2) enriched for RNA transcripts encoded by evolutionarily constrained genes, (3) depleted of RNA transcripts regulated by common DNA sequence variants, and (4) enriched for RNA transcripts implicated in higher-order brain functions by human population genetic studies. In PFC excitatory neurons of living study participants, higher expression of TPAWN tracked with higher expression of RNA transcripts that in rodent PFC samples are markers of excitatory neurons that connect the PFC to deep brain structures. TPAWN was reproduced by RNA transcript expression patterns differentiating living PFC samples from postmortem PFC samples, and significant differences between living and postmortem PFC samples were additionally observed with respect to (1) the expression of most primary RNA transcripts, mature RNA transcripts, and proteins, (2) the splicing of most primary RNA transcripts into mature RNA transcripts, (3) the patterns of co-expression between RNA transcripts and proteins (this result is presented in the ***Supplementary Information***), and (4) the effects of some DNA sequence variants on RNA transcript and protein expression. In the remainder of this section, two contributions the study makes to human brain research are discussed and limitations of the study with respect to its ability to achieve the main study objective are stated.

One contribution made by the study to human brain research is the identification of a transcriptional program associated with neurotransmission in the brain of living humans (i.e., TPAWN). The first study to perform neurotransmission DE by pairing RNA sequencing of human brain tissue samples with intracranial neurotransmission recordings from the same individuals was the study by Berto *et al.* that identified the iEEG DEFs used in this report^19^. By applying a similar study design to two LBP cohorts, the current study integrated three different types of neurotransmission DE signatures (i.e., FSCV DE signatures; MER DE signatures; iEEG DE signatures) derived from a total 146 neurosurgical procedures (FSCV DE analyses N = 15, MER DE analyses N = 115; iEEG DE analyses N = 16) to identify TPAWN. While the three types of neurotransmission DE analyses were of the same general experimental design (i.e., pairing RNA sequencing of brain tissue samples with intracranial neurotransmission recordings from the same individuals), the experimental designs also differed from one another in eight key ways. First, each type of neurotransmission DE analysis used a different intracranial recording technology (i.e., FSCV, MER, or iEEG). Second, for iEEG DE analyses, the brain region recorded and the brain region sampled were the same (i.e., the MTG), whereas for the FSCV and MER DE analyses the brain region recorded (for FSCV DE analyses, the SNr; for MER DE analyses, the STN and GPi) differed from the brain region sampled (i.e., the PFC). Third, the brain region sampled for the iEEG DE analyses differed from the brain region samples for the FSCV and MER DE analyses. Fourth, different brain regions were recorded for each type of neurotransmission DE analysis. Fifth, the timing of the brain recording and sampling relative to one another for the FSCV and MER DE analyses (i.e., biopsy followed 10 minutes later by recording) differed from the timing of the brain recording and sampling relative to one another for the iEEG DE analyses (i.e., recording followed days later by biopsy). Sixth, different metrics were used to represent neurotransmission for each of the three types of neurotransmission DE analyses (i.e., for FSCV DE analyses, the neurotransmitter association with offer size; for MER DE analyses, the aperiodic exponent; for iEEG DE analyses, waveforms). Seventh, for the FSCV and iEEG DE analyses, study participants engaged in tasks that engaged higher-order brain function during the recordings, whereas for the MER DE analyses study participants were at rest during the recordings. Eigth, for the FSCV and MER DE analyses the study participants were individuals undergoing DBS electrode implantation to treat PD and other conditions that are indications for DBS, whereas for the iEEG DE analyses the study participants were individuals undergoing surgery to treat epilepsy. These eight key differences in experimental design are a strength of this study: the convergence of the neurotransmission DE signatures on TPAWN despite these experimental design differences speaks to the robustness and generalizability of the signal. However, these experimental design differences also render interpretation of the results difficult. For example, the precise role of a given TPAWN gene in neurotransmission cannot be determined from this study. To the extent that it is feasible, future larger studies of diverse neurosurgical cohorts should aim to systematically pair molecular measurements from across the brain to neurotransmission measurements from across the brain to gain a deeper understanding of the observations made here. Such knowledge could facilitate the development of therapeutics that target neural circuit activities in a precise manner to treat neurological and mental illnesses.

A second contribution the study makes to medical research is that it expands knowledge of how living and postmortem human brain tissues differ at the molecular level. Comparing living and postmortem human brain tissues can be viewed as a scalable approach for characterizing the molecular foundations of brain tissue function given that (1) the observed differences converged with the neurotransmission DE signatures and (2) performing studies that pair brain sampling with intracranial recordings in humans may be difficult to implement in at scale in the absence of significant resources. Knowledge of how living and postmortem human brain tissues differ at the molecular level is also of value for informing the design of studies that only utilize postmortem tissue. The LBP studies reported by Liharska *et al.*^14^, Vornholt *et al.*^17^, and Rodriguez de los Santos *et al.*^49^ characterized the differences in RNA transcript expression between living and postmortem human PFC tissues. By applying different analytic strategies to the same bulk RNA-seq data analyzed in those studies, as well as by integrating that data with genomic and proteomic data introduced here, the current report shows that differences exist between living and postmortem human brain samples with respect to (1) the expression levels of most primary RNA transcripts, mature RNA transcripts, and proteins, (2) the splicing of most primary RNA transcripts into mature RNA transcripts, (3) the patterns of co-expression between RNA transcripts and proteins, and (4) the effects of some DNA sequence variants on RNA transcript and protein expression. In addition to finding many molecular differences between living and postmortem samples, this study also found some key similarities. For example, the set of CRFs identified appears to be robust to the LIV-PM status of the samples used in the analysis. Therefore, as has been noted in earlier LBP reports, depending on the research question of interest, the molecular differences that exist between living and postmortem human brain tissues may or may not matter, and future work should aim to determine which research objectives can be adequately achieved using only postmortem human brain tissue. The current report demonstrates one example of a research objective (i.e., to characterize the multiomic foundations of functional brain tissue) that required both living and postmortem samples to achieve.

The study had several limitations with respect to its ability to achieve the main study objective. Limitations for the neurotransmission DE analyses, discussed in more detail above, include lack of power and inconsistently designed neurotransmission DE experiments. A limitation of the LIV-PM DE analyses is that the ability of these analyses to achieve the main study objective rests upon the assumption that molecular differences between living and postmortem samples are representative of molecular differences between PFC tissue from individuals with intact higher-order brain functions and PFC tissue from individuals lacking intact higher-order brain functions. In all likelihood, the functional capacity of the individual at time of PFC sampling is just one of many variables captured by the LIV-PM DE signatures. Concern regarding this limitation is mitigated by (1) the convergence between LIV-PM DE signatures and TPAWN and (2) the analyses in the ***Supplemental Information*** showing that the LIV-PM DE signatures are stable with respect to 12 potential confounding variables. Future studies in model systems (e.g., cell lines, rodents) may be of value for more clearly delineating all the variables that contribute to the LIV-PM DE signatures, and the mechanisms by which these variables make their contributions. In particular, those studies should seek to establish the extent to which the procedures used to acquire and process PFC biopsies from living participants could impact RNA transcript and protein expression. Another limitation of the study design is that while RNA-seq and LC-MS are state-of-the-art technologies for characterizing the transcriptome and proteome in human tissues, respectively, these technologies remain limited in their ability to fully capture the multiomic foundations of tissue function. This is evident in some of the study results, such as the low same-gene RNA-protein correlations observed in both LIV samples and PM samples (this result is presented in the ***Supplementary Information***). When these technologies are succeeded by improved technologies in the future, studies should aim to refine the observations made here.

As noted at the outset of this report, an aim of medical research is to advance knowledge of how higher-order brain functions emerge from human brain tissue. Tools available to researchers who study the brain in humans to achieve this aim include tools for assessing higher-order brain functions (e.g., the ultimatum game), tools for recording brain activity (e.g., intracranial recordings), and tools for characterizing brain biology (e.g., RNA sequencing). Most applications of the tools available for characterizing human brain biology have been applied to samples obtained from individuals in the postmortem setting, when neurotransmission and higher-order brain function have ceased. As a result, the multiomic foundations of human brain functions are not being adequately studied. The current study illustrates how the scope of neuroscience questions addressed in human subjects can be broadened by studying biology in brain tissue obtained during neurosurgical procedures in conjunction with research activities that use tools for assessing higher-order brain functions and for recording brain activity. The findings from this report indicate that the multiomic foundations of human brain function likely involve tens of thousands of RNA transcripts and proteins interacting in highly complex ways, and to fully characterize these processes will require much larger studies of brain tissue samples obtained from living people in a safe and ethical manner.

## Data Availability

All data analyzed from the full LBP cohort are available via the AD Knowledge Portal (https://adknowledgeportal.org). The AD Knowledge Portal is a platform for accessing data, analyses, and tools generated by the Accelerating Medicines Partnership (AMP-AD) Target Discovery Program and other National Institute on Aging (NIA)-supported programs to enable open-science practices and accelerate translational learning. The data, analyses and tools are shared early in the research cycle without a publication embargo on secondary use. Data is available for general research use according to the following requirements for data access and data attribution (https://adknowledgeportal.org/DataAccess/Instructions). For access to content described in this manuscript see: https://www.synapse.org/#!Synapse:syn51622714/datasets/

## Acknowledgments

The Living Brain Project study participants are commended for their important role in science. The following centers, programs, departments and institutes within the Icahn School of Medicine at Mount Sinai supported the work: Charles Bronfman Institute of Personalized Medicine; Department of Neurosurgery; Department of Genetics and Genomic Sciences; Department of Psychiatry; Department of Neuroscience; Department of Medicine; Friedman Brain Institute; Center for Neuromodulation; Center for Computational Psychiatry; Nash Family Center for Advanced Circuit Therapeutics. The following centers, programs, departments and institutes within the Virginia Tech supported the work: Fralin Biomedical Research Institute; Department of Physics. The following centers, programs, departments and institutes within the Aarhus University supported the work: Center of Functionally Integrative Neuroscience. The following centers, programs, departments and institutes within the University College London supported the work: Wellcome Centre for Human Neuroimaging. The following centers, programs, departments and institutes within Oxford supported the work: Department of Experimental Psychology. The following centers, programs, departments and institutes within Rice University supported the work: Department of Statistics. The following centers, programs, departments and institutes within Wake Forest supported the work: Department of Translational Neuroscience; Department of Neurosurgery. Postmortem samples from Harvard Brain Tissue Resource Center and University of Miami Brain Endowment Brain were acquired under the National Institutes of Health NeuroBioBank request number 543. Postmortem samples from the New York Brain Bank of Columbia University were acquired under request number 1962. Thank you to Dr. Xiaosi Gu and her team (Arianna Neal Davis, Ofer Perl, Qixiu Fu, Matthew Heflin) for helping with the voltammetry data collection.

## Author contributions

Conceptualization: AWC, NBD, BHK

Study design: AWC, NBD, BHK, IS, PRM

Data quality control and analyses: AWC, NBD, DK, LEL, EV, AV, AL, RCT, TL, WM, NB, AA, JH, CF, SRB, DB, LSB, TT, JPW, MV, BHA, MSB, KTT, IS, PRM

Writing – original draft: AWC, LEL

Writing– review and editing: All authors

Funding acquisition: AWC, EES, DMR, GNN, NRN, MAK,

Participant recruitment and sample acquisition: AWC, BHK, AH, LW, KZ, HS, LML, BS, VC, PK, EM, MF, JS

Sample processing and multiomic data generation: AWC, YJP, EC, JSJ, PS, RS, SQ, GMM, NMC, MKR, NRN, MAK

---

**Supplementary Figure 1.**
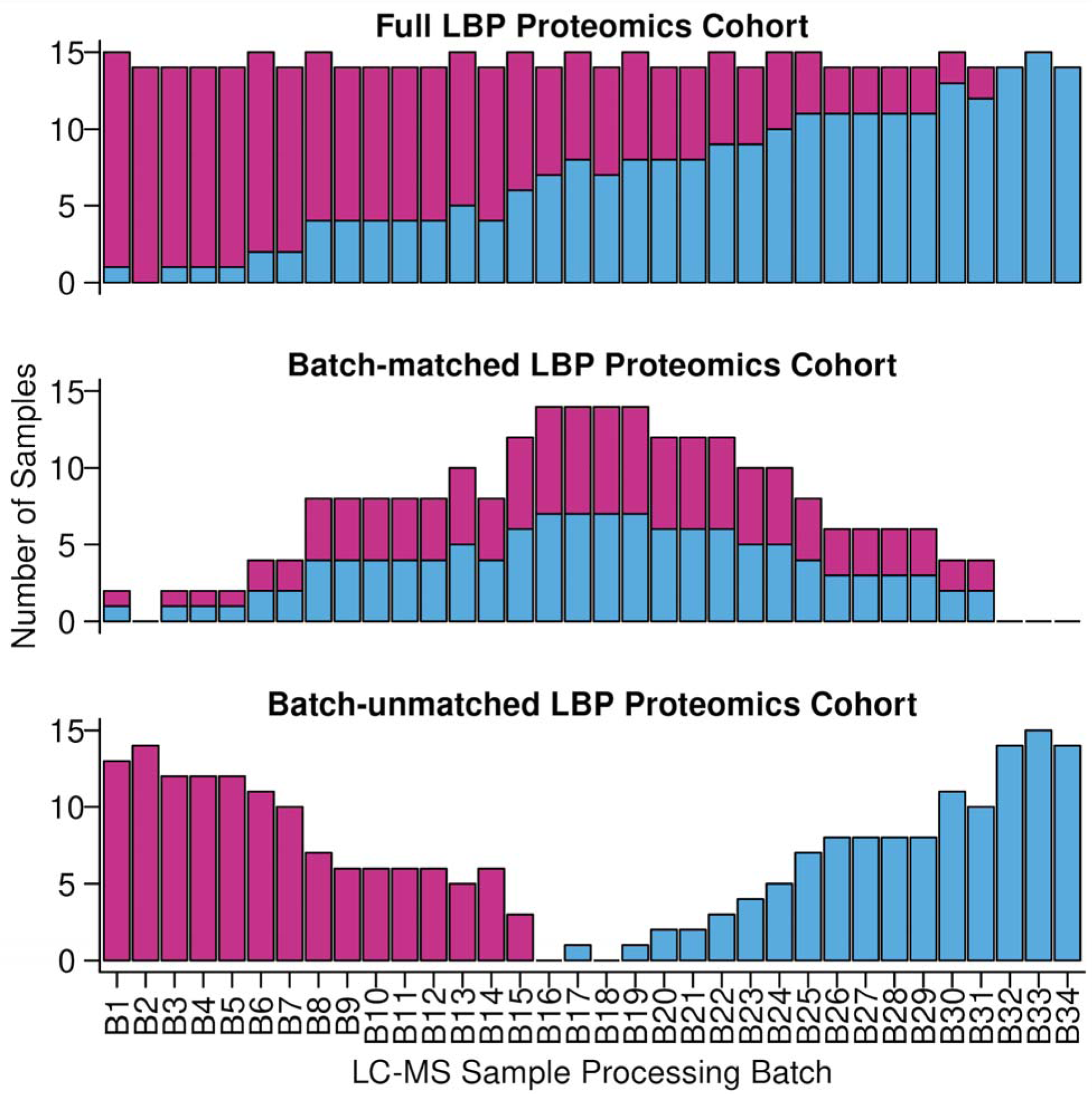
Barplots showing the number of LIV samples and PM samples (y-axis) in each experimental batch of bulk LC-MS data (x-axis). The top plot shows counts for the full set of PFC samples in the protein LIV-PM DE analysis. The middle plot shows counts for the set of PFC samples in the “batch-matched” protein LIV-PM DE analysis. The bottom plot shows counts for the set of PFC samples in the “batch-unmatched” protein LIV-PM DE analysis. See ***Supplementary Information*** for descriptions of how the batch-matched and batch-unmatched sets of samples were defined.

**Supplementary Figure 2.**
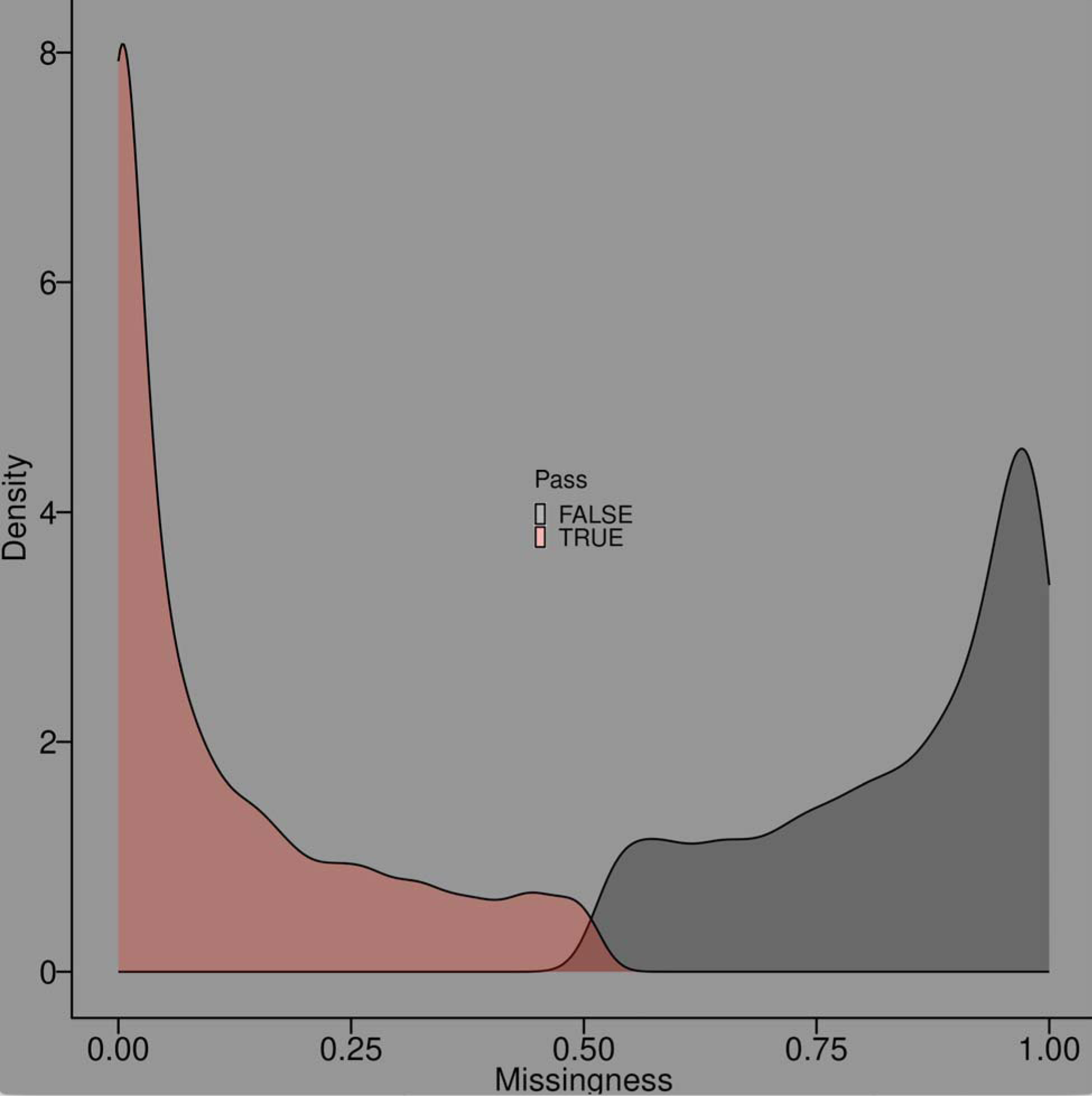
Density plot showing the distribution of missingness rates for the 10,515 proteins quantified in the bulk LC-MS data prior to quality control. The colors indicate whether the proteins in the distribution were retained for analysis after quality control (red) or were removed from analysis during quality control procedures (gray).

**Supplementary Figure 3.**
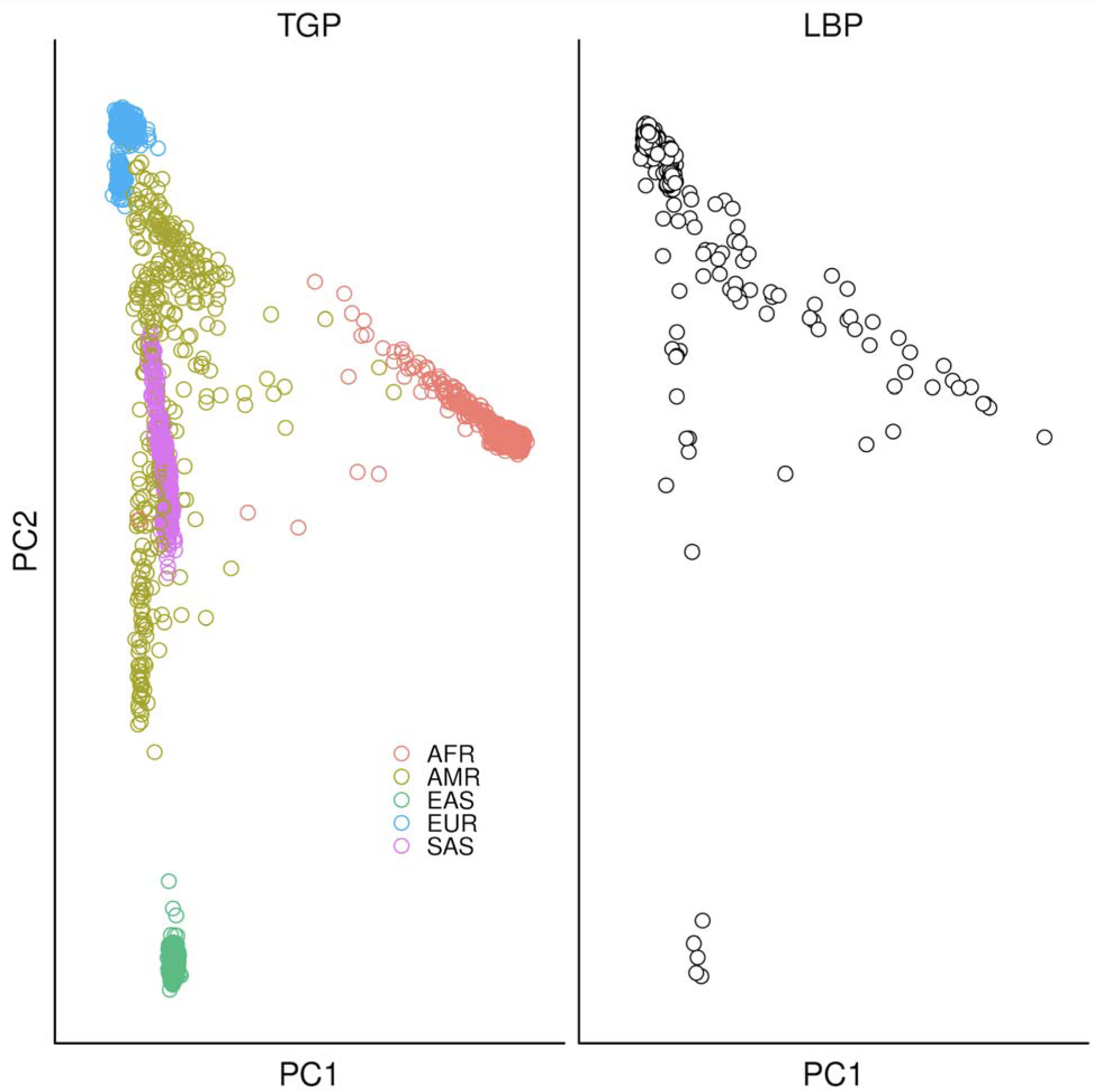
Scatter plots showing top two genetic principal components for individuals in the Thousand Genomes Project (TGP; left panel) and the LBP for the current report (right panel). TGP data is provided as a reference to show the genetic ancestry breakdown of the LBP cohort.

**Supplementary Figure 4.**
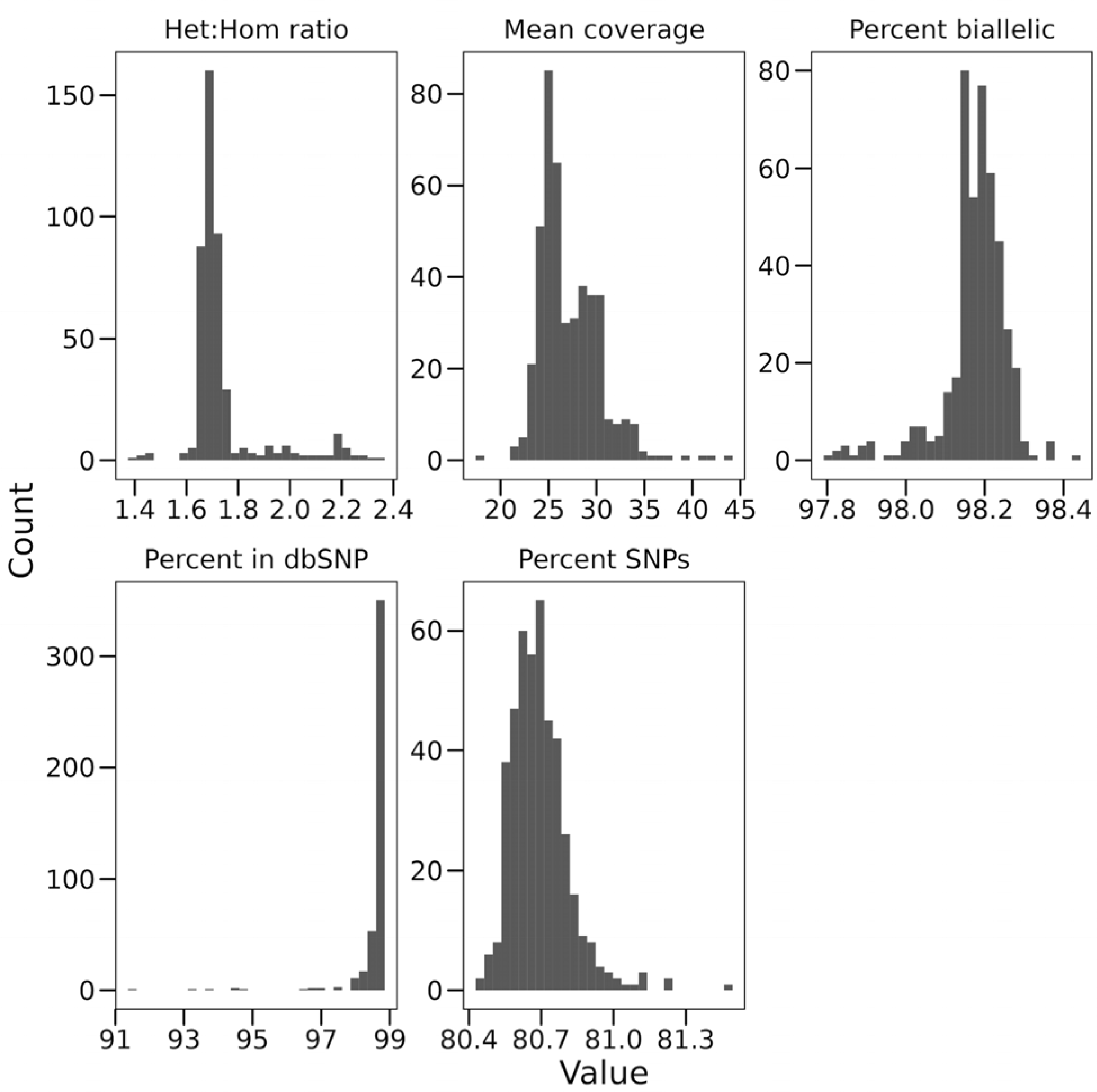
Each panel is a histogram showing the distribution of quality metric values for the LBP WGS data after quality control procedures. The quality metric is indicated in the panel title.

**Supplementary Figure 5.**
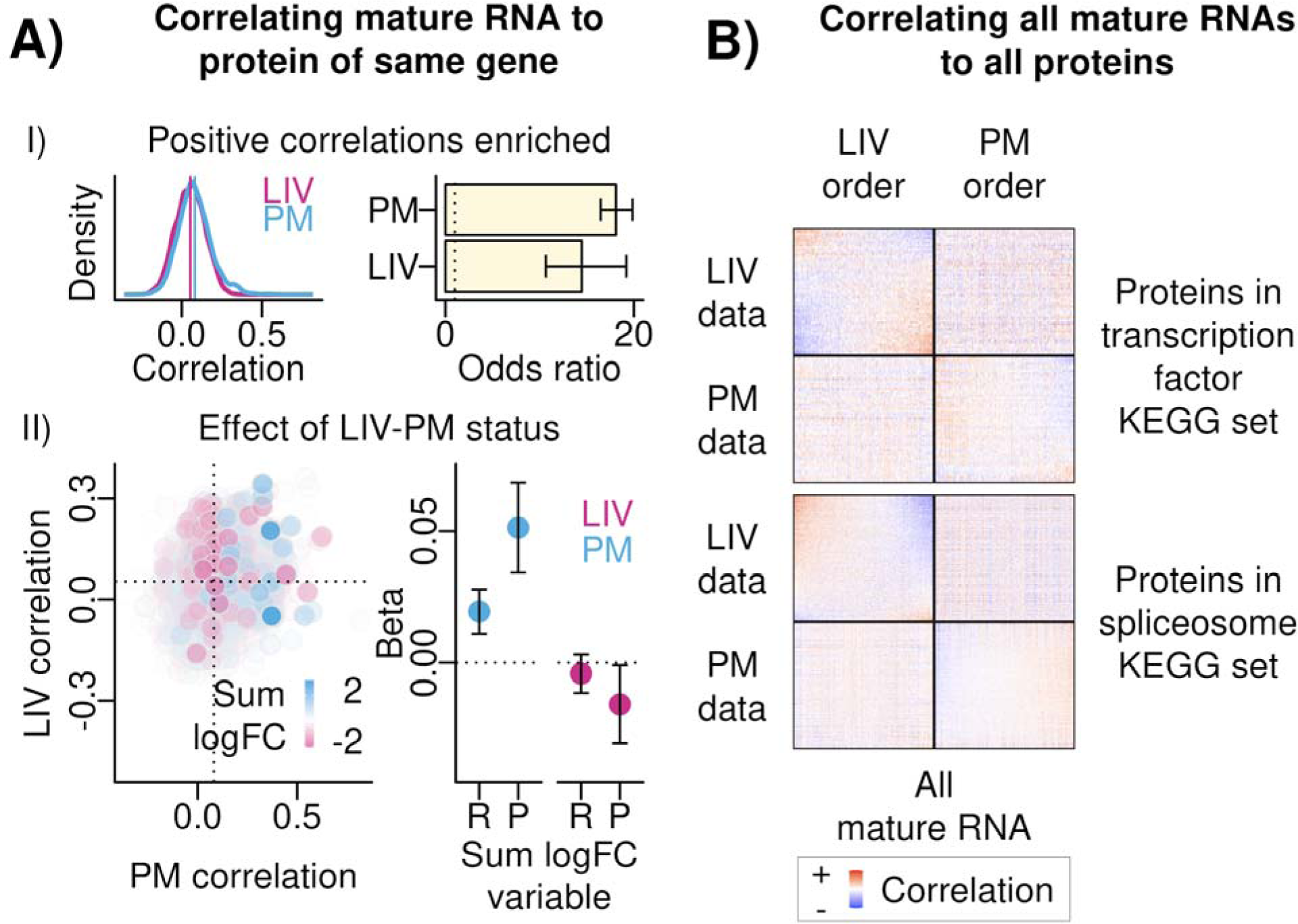
(A) Correlating mature RNA to protein of same gene. (I) *Positive Correlations Enriched.* Left – density plots of the same-gene RNA-protein correlations in LIV samples (pink) and PM samples (blue). Right – Bar plot showing the results of tests of whether the same-gene RNA-protein correlations were significant and positive more than expected by chance in LIV samples and PM samples (vertical axis). The summary statistic of this test is the odds ratio, presented on the horizontal axis. Error bars are the 95% confidence interval of the odds ratio from Fisher’s exact test, and error bars that do not cross the dotted vertical line intersecting the horizontal axis at 1 indicate statistical significance. (II) *Effect of LIV-PM status*. Left – scatter plot showing relationship of same-gene RNA-protein correlations in PM sample (x-axis) to same-gene RNA-protein correlations in LIV samples (y-axis). To color and shade the points, the mature RNA logFC and the protein logFC were summed; if the sum was greater than 0 the point was colored blue, if the sum was less than 0 the point was colored pink, and the shade of the points was set to reflect the absolute value of the sum. Right – To determine if LIV-PM status was driving same-gene mature RNA-protein correlations, betas (y-axis) and associated p-values were obtained using the following linear model applied separately to LIV samples and PM samples (x-axis): same-gene correlation coefficient ∼ Mature RNA LIV-PM logFC (resulting beta is “R” on x-axis) + Protein LIV-PM logFC (resulting beta is “P” on x-axis). <u>(B)</u> <u>Correlating all mature RNAs to all proteins.</u> For the two KEGG sets most enriched for differentially correlated RNA-protein pairs (transcription factors; spliceosome) the RNA-protein correlations are shown between the proteins in the set (heatmap rows) and all mature RNA transcripts (heatmap columns). Positive correlations are in red, negative correlations are in blue. “LIV Data” and “PM Data” describe the set of samples (i.e., LIV samples or PM samples) used to generate the correlations in the corresponding row of heatmaps. “LIV Order” and “PM Order” describe the data that was used to order the proteins and mature RNA transcripts in the corresponding column of heatmaps.

**Supplementary Figure 6.**
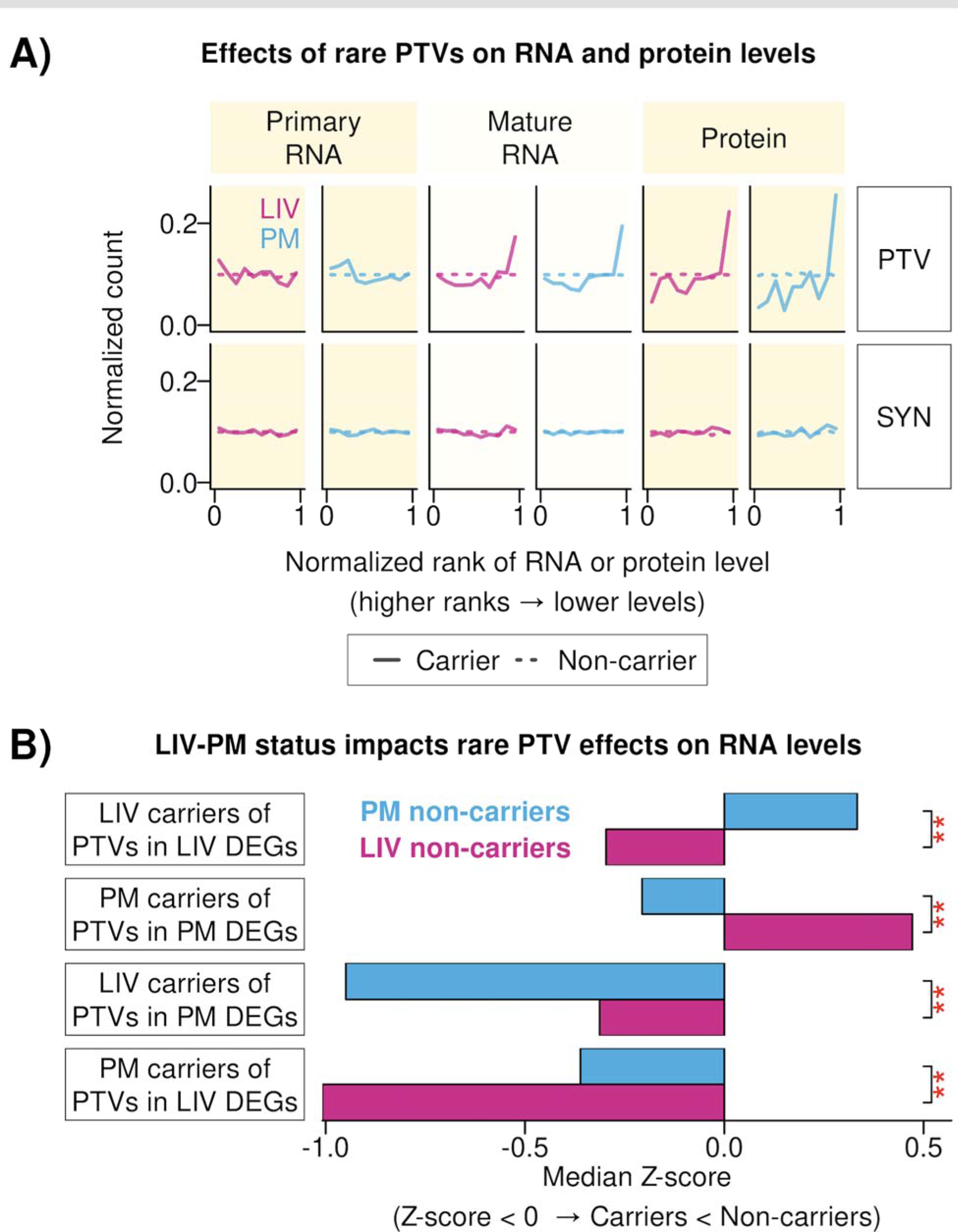
(A) Effects of rare PTVs on RNA and protein levels. To determine if LIV-PM status impacts the ability to detect the effects of rare PTVs on RNA transcript and protein expression, for primary RNA transcripts, mature RNA transcripts, and proteins, the following four-step procedure was performed separately in LIV samples (pink lines) and PM samples (blue lines): (1) for each RNA transcript and protein, expression levels were normalized into “expression ranks” (i.e., the rank of a sample in a cohort with respect to the expression level, divided by the number of samples in the cohort), such that (a) expression ranks ranged from 0 to 1 and (b) higher expression ranks reflected lower expression levels; (2) for each RNA transcript and protein, the expression rank of each sample was annotated as either a “carrier expression rank” (i.e., the sample was a carrier of a rare PTV in the gene coding for the RNA transcript or protein) or a “non-carrier expression rank” (i.e., the sample was not a carrier of a rare PTV in the gene coding for the RNA transcript or protein); (3) the second step of the procedure was repeated for rare synonymous variants instead of rare PTVs. The third step of the procedure was performed for control purposes, as there is no expectation that expression ranks should differ between carriers and non-carriers of rare synonymous variants. Each plot is a histogram of the resulting expression ranks (horizontal axis). The row labeled “PTV” shows the expression ranks for analyses of rare PTVs and the row labeled “SYN” shows the expression ranks for the analyses of rare synonymous variants. Plots with pink lines show the expression ranks from analyses of LIV samples and plots with blue lines show the expression ranks from analyses of PM samples. Solid lines represent the distribution of expression ranks in carriers of rare variants and dotted lines show the distribution of expression ranks in non-carriers. Each line connects ten data points, and the horizontal axis values of the data points start at 0.05 and are separated from one another by 0.10. Each point summarizes the distribution of expression rank values in the corresponding decile (e.g., the point at 0.05 summarizes the expression ranks for the 0.00 – 0.10 decile). For the analysis given by the histogram position in the plot grid, the line color, and the line type (e.g., primary RNA transcript, rare PTVs, LIV samples, rare variant carriers), the vertical axis value for each of the 10 connected points in the histogram was calculated by dividing the number of expression ranks in the corresponding decile by the total number of expression ranks in the analysis represented by the line. <u>(B) LIV-PM status impacts rare PTV</u> <u>effects on RNA levels.</u> For LIV DEFs and PM DEFs, the expression ranks in carriers of rare PTVs were compared to expression ranks of non-carriers, while varying the LIV-PM status of the non-carrier group. This resulted in 8 conditions, which are represented by the 8 bars in the bar plot: (1) LIV carriers of rare PTVs in LIV DEFs compared to (a) LIV non-carriers and (b) PM non-carriers; (2) PM carriers of rare PTVs in PM DEFs compared to (a) LIV non-carriers and (b) PM non-carriers; (3) LIV carriers of rare PTVs in PM DEFs compared to (a) LIV non-carriers and (b) PM non-carriers; (4) PM carriers of rare PTVs in LIV DEFs compared to (a) LIV non-carriers and (b) PM non-carriers. The colors of the bars in the bar plot indicate the LIV-PM status of the non-carrier group. For each of the 8 conditions, Z-scores were calculated for every DEF tested under the condition as the difference between the expression ranks of rare PTV carriers and the expression ranks of non-carriers. The median Z-scores of the DEFs tested under each condition are plotted on the horizontal axis. Negative Z-scores occurred when the carrier(s) had lower expression than non-carriers. The difference in Z-score distributions were then calculated between the LIV non-carrier and PM non-carrier versions of the condition using a K-S test, and the significance of those tests are indicated by the red asterisks.

## Methods

### Ethics statement

All human subjects research was carried out under STUDY-13-00415 of the Human Research Protection Program at the Icahn School of Medicine at Mount Sinai. Research participants in the living cohort provided informed consent for sample collection, genomic profiling, clinical data extraction from medical records, and public sharing of de-identified data. A subset of the participants additionally consented to the FSCV recording component of the study, which consisted of FSCV recordings and the ultimatum game.

### Living Brain Project cohort

All of the individuals and PFC samples studied in the current report were first introduced in earlier LBP reports^14^. In the methods sections of those reports can be found descriptions of the living cohort, the postmortem cohort, PFC sample collection procedures, and clinical data collection (i.e., anesthesia given to the living cohort, PD symptom severity in the living cohort, and dopamine replacement therapy in the living cohort).

### Fast-scan cyclic voltammetry (FSCV) recordings

FSCV recordings were made during 15 DBS surgeries. The PFC samples obtained during these 15 surgeries were not part of the 289 LIV samples processed for bulk RNA-seq data. The procedures used for making the FSCV recordings are fully described in Batten *et al.*^16^ and summarized in this section.

#### Experimental setup

FSCV recordings were made while study participants played 60 rounds of the ultimatum game^28,29^. For 6 of the 9 participants in this component of the LBP, the ultimatum game was performed twice (i.e., in separate DBS surgeries) and for 3 participants it was performed once. Participants performed a short practice session both before and within a DBS surgery. During DBS surgery, study participants laid in a semi-upright condition and viewed a task presentation monitor at a distance of around 100 centimeters. They used a gamepad to submit their responses to the trial offers. The FSCV recordings were obtained approximately 10 minutes after PFC biopsies.

#### Description of the ultimatum game

In each round of the ultimatum game, the participant must decide whether to accept or reject a monetary offer made by an avatar (the “proposer”). The proposer offers a split of $20 to the participant, and the offer is allowed to range from $1 to $9. If the study participant accepts the offer, they get the offered amount and the proposer gets the remaining amount (i.e., the difference between $20 and the offer). If the participant rejects the offer, neither the participant nor the proposer receives any money.

#### FSCV recording technology

Approximately 10 minutes after PFC biopsy, electrochemical recordings were made from the substantia nigra pars reticulata (SNr) using a custom-made carbon-fiber (CF) electrode^25^. From these recordings, the estimates of dopamine, serotonin, and norepinephrine fluctuations were made using deep convolutional neural networks that were trained and tested on *in vitro* data that consisted of 64 datasets collected by exposing 64 CF electrodes to varying concentrations of dopamine, serotonin and norepinephrine in a saline solution^16,50^.

#### Deriving FSCV metrics used in differential expression analyses

For each neurotransmitter, activity during a proposed offer period was measured as the area under the curve (AUC) of the neurotransmitter measurements made during the offer period. The offer period was defined as the one second following the moment the offer was first shown to the subject. Neurotransmitter measurements were made at 10 points during this period spaced 0.1 seconds apart, and the AUC value was calculated as the sum of these measurements. The measurement at time 0 was subtracted out of each measurement before the sum was calculated. Since 60 offers were made during the game, 60 AUC values for each neurotransmitter resulted from this process. Both the AUC values and offer amounts were then transformed into z-scores, which were then input into a linear regression model using the following formula to calculate beta coefficients between neurotransmitter AUC values and offer amount values. These analyses were performed using the Statistics Toolbox in MATLAB R2023a.

<u>Formula 1 (linear model with fixed effects):</u>

*Neurotransmitter AUC ∼ Offer amount + Residuals*

### Microelectrode recordings (MERs)

Intracranial MERs are routinely obtained for clinical purposes from awake patients during DBS surgery to pinpoint where to place the DBS electrode. For this report, MERs were analyzed from a subset of the DBS surgeries where bulk RNA-seq had been performed on the PFC biopsy obtained during the surgery. MERs were obtained approximately 10 minutes following PFC biopsies using the Guideline 4000 (FHC, Inc. Bowdoin, ME) and a tungsten monopolar microelectrode (22675Z, FHC, Inc. Bowdoin, ME) with a 44000 Hertz (Hz) sampling frequency and analyzed offline.

MERs were retained for analysis if (1) the MER was taken in the STN or GPi, as determined by the distance between the microelectrode and the target (STN: 0-5 mm, GPi: 0-6 mm); (2) the duration of the MER was at least 5 seconds. To exclude artifacts and the need to include an ‘aperiodic knee’ in subsequent analysis steps, a finite input response bandpass filter of 2-50 Hz was applied to the local field potential (LFP) time series data derived from the MERs using the filter_signal() function within the NeuroDSP Python package (v2.2.1). The output of this filter was then down sampled to 1000 Hz using the signal.resample() within the Scipy Python package (v1.12.0). Power spectrum densities (PSD) within the frequency range of 2-50 Hz were then calculated with the Welch method for each MER using the compute_spectrum() function within the NeuroDSP Python package.

Spectral parameterization was then applied to extract timescales from PSDs using the FOOOF() function from the FOOOF Python package (v1.1.0)^51^, which decomposes log-power spectra into periodic components and the aperiodic exponent. Periodic components are modeled as Gaussians, and the aperiodic exponent is modeled as a generalized Lorentzian function centered at 0 Hz. This approach is preferred for analyzing neural signals wherein a strong 1/f power law is present. This allows for oscillatory, or periodic, components to be easily accounted for and ignored in the frequency domain when inferring the aperiodic exponent of the PSD^51^. Each PSD was fit across the FOOOF algorithm with settings (peak_width_limitsL=L[2,7], max_n_peaksL=L3, min_peak_heightL=L0.05, peak_thresholdL=L2, aperiodic_modeL=L‘fixed’) across the frequency range 5–45 Hz. This provided an aperiodic exponent value that reveals baseline power across frequencies (1/f), reflecting the balance of neuronal excitation and inhibition within each LFP time series. Recordings with a FOOOF model fit of R^2^ < 0.8 and aperiodic exponent < 0 were excluded from analyses to avoid the use of data from poor quality recordings.

### Bulk RNA-seq

In this section, the methods used to perform bulk RNA-seq and prepare the resulting bulk RNA-seq data for analysis are presented in full. Towards this end, four “pipelines” (where a pipeline is defined as a sequential series of steps that together achieve a specific goal) were implemented:

1. RNA Sequencing Pipeline: the goal of this pipeline is to generate the bulk RNA-seq data for analysis.
2. Confounder Identification Pipeline: the goal of this pipeline was to identify unwanted drivers of variance in RNA transcript expression to include as covariates in bulk RNA-seq data analyses.
3. Primary and Mature RNA Quantification Pipeline: the goal of this pipeline was to concurrently quantify the levels of primary and mature forms of each RNA transcript detected in the bulk RNA-seq data.
4. Intron Usage Quantification Pipeline: the goal of this pipeline was to quantify intron usage in bulk RNA-seq data.

#### RNA Sequencing Pipeline

The methods used to perform RNA extraction, RNA sequencing, cell type deconvolution, and data quality control for the bulk RNA-seq data analyzed in the current report are introduced and fully described in Liharska *et al.*^14^ and briefly summarized here. Approximately 5-10 milligrams of each sample was used for RNA extraction. Extraction was generally performed in batches of 12 samples using the RNeasy Kit (Qiagen, Hilden, Germany), mostly according to manufacturer instructions. Only specimens with an RNA integrity number (RIN) greater than 4.0 were sent for RNA sequencing. Preparation of cDNA libraries and RNA sequencing were performed at Sema4 (Stamford, CT). Libraries of cDNA were prepared using the TruSeq Stranded Total RNA with Ribo-Zero Globin Kit (Illumina, San Diego, CA; Catalog Number 20020613). RNA sequencing was performed on the NovaSeq 6000 System (Illumina, San Diego, CA). Base calls were made from data emitted by the clusters within the S4 flow cell and organized into sequencing reads stored in FASTQ files using Illumina’s bcl2fastq software (v1.8.4).

#### Confounder Identification Pipeline Summary

The methods used to identify confounders to account for in the bulk RNA-seq data analyzed in the current report are introduced and fully described in Liharska *et al.*^14^ and briefly summarized here. Unaligned reads were aligned to the GRCh38 primary assembly with Gencode gene annotation v30 using STAR (v2.7.2a)^52,53^. Aligned reads were sorted using samtools (v1.13)^54^ and duplicate reads were marked using Picard Tools (v2.20.1). Unwanted drivers of variance in RNA transcript expression to include as covariates in bulk RNA-seq data analyses were identified using an iterative pipeline that considered over one hundred technical variables.

#### Primary and Mature RNA Quantification Pipeline

FASTA and General Transfer Format (GTF) source files for the human genome GRCh38 primary assembly with Gencode gene annotations v41 were obtained from the Gencode website. (https://ftp.ebi.ac.uk/pub/databases/gencode/Gencode_human/release_41/)^53^. The GTF file was input into the getFeatureRanges() function of the eisaR R package (v1.8.0) with the featureType parameter set to “spliced” (which corresponds to mature RNA) and “unspliced” (which corresponds to primary RNA) and default settings used for the other parameters^55^. With these settings, the getFeatureRanges() function parses the input GTF to return an object that for each transcript in the GTF annotated in the GTF as having >=1 exon there are two entries: one that provides a genomic range for each exon (i.e., its start and stop positions) and one that contains a single genomic range for the transcript that is inclusive of all the exon ranges as well as the genomic ranges separating the exon ranges (i.e., the intronic ranges). The getTx2Gene() of the eisaR R package was then applied to this object to create a file linking transcript identifiers to gene identifiers that was then used in subsequent steps described below. The GRCh38 FASTA file was then read into R using the readDNAStringSet() function of the Biostrings R package (v2.60.1). The resulting object, along with the object output by the getFeatureRanges() function, were then input into the extractTranscriptSeqs() function of the GenomicFeatures R package (v1.44.0). This function iterates over each entry in the object output by the getFeatureRanges() function and extracts the genomic sequence corresponding to the ranges in the entry. For entries that correspond to mature RNA, the resulting sequence is a single string of the combined exonic sequences. For entries that correspond to primary RNA, the resulting string contains the full transcript sequence (i.e., exons and introns). The resulting set of sequences were then saved as a FASTA file using the writeXStringSet() function of the Biostrings R package. The newly created FASTA was then input into the index function of the salmon unix package (v1.9.0) along with the full reference FASTA from which it was created. The two FASTA files were supplied as the “--transcripts” argument to the index function along with the “--gencode” flag (specifying the FASTA files are in Gencode format) and a list of all sequence names in the full reference FASTA (e.g., chromosome names) to the “--decoys” argument^56^ and otherwise default parameters^56^. The output of this step was a salmon index (“SIDX”) file.

For each of the 518 samples with bulk RNA-seq data the salmon quant command was run giving as input the FASTQ files containing the RNA sequencing reads for the sample (the “--mates1” and “--mates2” arguments), the SIDX file (to the “--index” parameter), “--libType” set to ISR, and otherwise default settings. From the resulting set of quantification files gene-level counts were derived for each sample using the tximport() function of the tximport R package (v1.20.0) with the “type” parameter set to “salmon” and the “countsFromAbundance” parameter set to “lengthScaledTPM” and the “tx2gene” parameter set to a path to a file linking transcript to gene identifiers created as described above during the preparation of the SIDX file^57,58^. Separate counts matrices were created for primary RNA (38,387 features) and mature RNA features (61,436 features; 38,344 overlapping with primary RNA). Lowly expressed RNA features were filtered out by calculating the log of the mean of the gene-level TPM (calculated by summing the transcript-level TPM values output by salmon quant). Any gene with a value less than negative five was removed, leaving 22,955 primary RNA and 30,099 mature RNA features for analysis. The counts of these features output by tximport were normalized using the voomWithDreamWeights function of the dream software within the variancePartition R package (v1.20.0) because two samples per individual were available for some individuals^59,60^.

#### Intron Usage Quantification Pipeline

The “junction extract” command of the regtools unix package (v0.5.1) was run to quantify the usage of each intron based on CIGAR strings from the BAM files generated as described above^61^. Default settings were used with the exception that the minimum intron length (“-m”) was set to 50 and the strand specificity (“-s”) was set to 2.

The file output by regtools, which contains one row per intron and the corresponding intron usage for the sample, was then used as the input to LeafCutter (v0.2.9) in order to identify intron clusters^41^. The purpose of the intron clustering step in LeafCutter is to represent as a unit different forms of intronic excisions that occur on the same region. LeafCutter defines intron clusters in a manner such that every intron in the cluster must share a start or stop position with at least one other intron in the cluster. When represented as a graph where each intron is an edge and intron start and stop positions are vertices, an intron cluster is a connected graph (i.e., there is a path between all vertex pairs). Intron clusters were identified using the LeafCutter “leafcutter_cluster_regtools.py” script, which was given as input the regtools output for all samples together (N = 518) and settings such that 50 split reads were required to support the definition of an intron cluster and introns spanning up to 500,000 base pairs in size were considered. The output of this script was a matrix of counts with one row per intron (i.e., multiple rows per intron cluster) and one column per sample.

A series of filtering steps were then applied to either introns or intron clusters in the output of the LeafCutter intron clustering step, adapted from a previous study on human aging^62^. For each intron, the percentage of samples with 0 counts was calculated and any intron with >=25% of the samples having counts of 0 were flagged for removal. For each intron cluster, the contribution of each intron to the cluster was calculated as follows: the counts for the intron were calculated by summing the counts for the intron across all samples; the counts for the intron cluster were calculated by summing the counts for all the introns in the cluster; the contribution of the intron to the cluster was the calculated as the intron counts divided by the intron cluster counts. Any intron with a contribution less than 5% was flagged for removal. After applying these intron filters, each intron cluster in the output was then reassessed to determine if the criteria used by LeafCutter to define intron clusters was still fulfilled - in other words, to determine that the cluster still was a connected graph when represented as a graph with introns as edges and intron start and stop positions as vertices. For a given intron cluster, there are three possible outcomes to this reassessment: (1) the intron cluster is found to have a single edge, in which case it is removed; (2) the intron cluster remains a connected graph, in which case it is retained; (3) the intron cluster is found to have multiple introns but is no longer a connected graph. In the event of the latter, the intron cluster was decomposed into subgraphs of connected components. If all subgraphs have a single intron, the intron cluster is removed. If any subgraphs have greater than one intron, a “pruning” procedure was performed on the intron cluster. The pruning procedure consists of decomposing the intron cluster to its connected subgraphs and retaining the subgraphs with the most introns (i.e., if there are three subgraphs and they have 2, 2, and 1 intron, the subgraphs with 2 introns will be both be retained and the subgraph with 1 intron will be pruned). If more than one subgraph is retained by this procedure, those subgraphs are split and considered separate intron clusters moving forward. After the pruning step, the number of introns in each intron cluster remaining was calculated and only intron clusters with 2-10 introns were retained.

### Bulk liquid chromatography-mass spectrometry (LC-MS)

#### Protein extraction and digestion

PFC samples were lysed with a lysis buffer containing 8M Urea, 50mM Tris-HCl pH 8.0, 1% Protease and Phosphatase Inhibitor Cocktail, and Optima LC-MS Water. Lysis buffer (200μL) was added to each sample and mixed by pipetting up and down, and then the whole sample was immediately transferred out of the sample vial and into a 1.5mL Eppendorf tube. Each sample was sonicated with four three-second pulses at 20% amplification to fully lyse the cells. Sonicated samples were centrifuged at 17,000 x g for 10 minutes, and the supernatant was then used in the Bradford Assay to determine the protein concentration. Proteins were reduced with 10mM Tris(2-carboxyethyl) Phosphine (TCEP) and alkylated with 18.75mM iodoacetamide Samples were diluted 1:5 with deionized water and digested with sequencing grade modified trypsin (Promega, Madison, USA; Catalog Number V5111) at a 1:50 enzyme-to-substrate ratio.

#### Peptide labeling

TMT (Tandem Mass Tag) reagents (16-plex; one “plex” labels all peptides from one sample so that peptides from multiple samples can be pooled together into a “multiplex”) were used to label peptides from all samples in 34 batches of pooled peptides (from 14-15 samples per batch), allowing relative quantitation (i.e., estimation of relative quantity) (Thermo Fisher Scientific, Waltham, USA; Catalog Number A44520). For tissue samples, a reference sample was created by pooling an aliquot of peptides from each individual sample. Peptides (20 µg) from each of the samples were dissolved in 20 µL of 200 mM triethylammonium bicarbonate (TEAB), pH 8.5 solution, and mixed with 20 µL of TMT reagent that was freshly dissolved in 256 µL of anhydrous acetonitrile, LC-MS grade. Channel 126 was used for labeling the pooled reference sample in all matrices analyzed in this study. After 4 hours of incubation at room tempterature, the reaction was quenched by adding 8 µL 5% hydroxylamine. Peptides labeled by different TMT reagents were then mixed, dried and desalted on C18 Spin columns. Desalted peptides were dried in a vacuum centrifuge and stored at −20°C until LC-MS. For technical reasons, it was not possible for each batch of pooled peptides to be comprised of equal numbers of LIV samples and PM samples.

#### Peptide fractionation

The newly formed batches were fractionated using basic reversed-phase liquid chromatography. Approximately 160 μg of 16-plex TMT labeled sample was first purified on C18 column, and then separated on a reversed phase Zorbax extend-C-18 column (4.6 × 100 mm column containing 1.8-um particles; Agilent, Santa Clara USA) using an Agilent 1200 Infinity HPLC System (Agilent, Santa Clara, USA). The solvent A consisted of 10 mM ammonium formate, pH 10.0. Solvent B consisted of 10 mM ammonium formate, pH 10, 90% acetonitrile as mobile phase. The separation gradient was set as follows: 2% B for 10 min, from 2 to 16% B for 10 min, from 16 to 40% B for 65 min, from 45 to 95% B for 5 min, and 95% B for 15 min. A total of 96 fractions were collected into a 96 well plate in a time-based mode. These fractions were then concatenated into 24 fractions by combining 4 fractions that are 24 fractions apart (i.e., combining fractions #1, #25, #49, and #73; #2, #26, #50, and #74; and so on). Each concatenated fraction was dried down in a Speed-Vac and re-suspended in 2% acetonitrile, 0.1% formic acid for LC-MS.

#### Mass spectrometry

LC-MS was performed using a Waters nanoAcquity LC (Waters, Milford, USA) system coupled to a Thermo Q Exactive Plus MS (Thermo Fisher Scientific, Waltham, USA). Peptides were separated in a 90 minute gradient from 5% B to 35% B. The eluting peptides were sprayed into the mass spectrometer using electrospray ionization and a data dependent Top 15 acquisition method was used to fragment candidate ions. Full MS survey scans were collected at a resolution of 35,000, scan range of 400-1800 Thompsons (Th; Th = Da/z), followed by MS/MS scans at a resolution of 35,000 with a 1.2 Th isolation window. Only ions with a +2 to +5 charge were considered for isolation and fragmentation. Data was searched using Proteome Discoverer 2.5 (Thermo Fisher Scientific, Waltham, USA) using SEQUEST algorithms and RefSeq database.

#### Data normalization, imputation, and quality control

Log2 transformed sample-to-reference ratio of MS2 (log2-ratio) intensity was produced for proteomics from 510 samples. In total 10,515 proteins were identified from the experiment, among which 2,310 were completely observed among all samples, and the overall missing rate was 39.7%. To preprocess the log2-ratio intensity data matrix, the procedure described in Wang *et al.*, 2021 was followed^63^. To remove the technical variation among the sample distribution globally, sample median alignment to log2-ratio intensity matrix was performed. For each protein, an “inter-TMT-multiplex t-test” was performed (between the log2-ratio of samples inside the TMT multiplex and the log2-ratio of samples outside of the TMT multiplex) to detect and remove outlier TMT multiplex values for the protein. For each protein, after double log-transformation, p-values of “inter-TMT-multiplex t-test” falling beyond four standard deviations from the median of the entire dataset were flagged as outliers, and the corresponding values of the specific protein for the respective TMT multiplex were labeled as “N/A” in the dataset. In total, outlier TMT multiplex values for six individual proteins were removed from the dataset (corresponding to six distinct TMT multiplexes for which all data for one respective protein was removed). Following outlier removal, the ComBat algorithm^63^ was applied to the dataset to correct for the effect of the TMT multiplex batch variable. In order to preserve the effect of LIV-PM status (which was correlated with TMT multiplex batch), the LIV-PM status variable was included in the ComBat model in addition to the TMT multiplex batch variable. Based on the resulting model values, only the calculated effect of TMT multiplex batch was regressed out of the protein expression data, while the calculated effect of LIV-PM status was preserved (i.e., protein expression was calculated as the sum of the effect of LIV-PM status and the residuals). Due to the ComBat algorithm’s requirement for complete data, KNN imputation was performed on the data prior to application of the ComBat algorithm using the impute R package^64^. After the imputed data was corrected using ComBat^65^, the missing data structure from before KNN imputation was restored. To formally impute the missing values, the DreamAI software was applied^66^. Imputation was done for the subset of 6,415 proteins that appeared in at least 50% of samples.

In order to identify unwanted drivers of variance to include as covariates in statistical models of protein expression, technical metrics and batch assignments characterizing the protein expression data were merged with the table of covariates characterizing the individuals (e.g., age, sex) and samples of the dataset. Covariates explaining the variance in protein expression between samples were then reviewed for potential inclusion in the statistical model of the data used for downstream analyses through an adaptation of an iterative procedure established in Liharska *et al.*^14^ for bulk RNA-seq data. First, LIV-PM status and individual ID were fit to a linear mixed model with individual ID as a random effect using the dream() function of the variancePartition R package, and residuals were calculated with the residuals() function of the base stats R package. Individual ID was accounted for as data from more than one sample per individual was available for some living participants. Accounting for the effect of LIV-PM status on the variance in protein expression at this step allowed for the potential effects of other covariates on the protein expression variance to subsequently be observed. Next, principal component analysis (PCA) was performed on the residual protein level matrix using the prcomp() function of the base stats R package and the canonical correlation between the variables in the covariate table and principal components (PCs) 1-5 of the residual RNA level data was calculated using the canCorPairs() function of variancePartition. Scatterplots of PCs (e.g., PC1 vs PC2) were generated and colored for each covariate as a visual aid in assessing the strength of the relationships between covariates and RNA levels. Canonical correlations and visual aids were reviewed and one covariate was considered for inclusion in downstream models. Due to the high correlation between LIV-PM status and TMT multiplex batch (canonical correlation = 0.59), and the fact that the data had already been corrected for TMT multiplex batch using the ComBat algorithm to remove the effect of TMT multiplex batch (and accordingly some partial effect of LIV-PM status also accounted for by TMT multiplex batch, as well as effects of any other correlated batch variables), the iterative procedure did not identify any additional covariates for inclusion in the statistical model of the data for downstream analyses. Samples were considered for removal for being outliers in the protein expression data. Outliers were defined as samples falling more than three standard deviations away from the centroid of PC1 and PC2 of the residual protein expression matrix after accounting for covariates selected using the above procedures. No samples were identified for removal.

### Immunohistochemistry

Six types of immunohistochemistry stains were performed on LIV samples and PM samples at the Icahn School of Medicine at Mount Sinai. For all six types of stains, tissue sections with a thickness of approximately 5 – 10 micrometers (μm) were formalin-fixed, paraffin-embedded, and baked on charged slides at 70 – 80°C for an average of 15 – 30 minutes.

#### Alpha-synuclein, amyloid, and tau staining

Chromogenic immunohistochemistry was performed on the BOND Rx Automated Research Stainer (Leica Biosystems, Wetzlar, Germany), as follows:

1. Heat-induced epitope retrieval was performed at a pH of 9 on the slides for 20 minutes
2. Slides were then incubated in the primary antibody for 30 minutes. The primary antibody used for Lewy body staining was mouse anti-alpha-synuclein (1:6000 antibody to diluent, LB509, MABN824MI, MilliporeSigma). The primary antibody used for amyloid staining was amyloid β (4G8) (1:4000 antibody to diluent, 4G8, BioLegend, Catalog # 800701). The primary antibody used for tau staining was Phospho-Tau (Ser202, Thr205) Monoclonal Antibody (AT8) (1:2000 antibody to diluent, AT8, Invitrogen, Catalog # MN1020).
3. The slides were then incubated for 20 minutes with the BOND Polymer Refine Detection kit (DS9800, Leica Biosystems, Wetzlar, Germany), which uses 3,3’-Diaminobenzidine (DAB) chromogen and hematoxylin counterstain to visualize the primary antibody in the tissue section.

Whole slide images were scanned at 40x magnification using the Versa 8 Scanner (Leica Biosystems, Wetzlar, Germany).

#### Luxol hematoxylin and eosin staining

Before undergoing sequential staining with Luxol Fast Blue and Hematoxylin and Eosin, slides were deparaffinized with the Leica ST5010 Autostainer XL by immersion XYLENE (3 stations, 4 minutes each), 100% EtOH alcohol (2 stations, 3 minutes each), 95% EtOH grade alcohol (2 stations, 3 minutes each), and wash (1 station, 3 minutes). Luxol Fast Blue staining was then done by hand, immersing slides in Luxol Fast Blue solution (1%) at 60°C for 1 to 2 hours with a tightly capped container then rinsing sequentially with 95% ethanol (1 to 3 dips), distilled water, Lithium Carbonate Solution (0.1%) (2 to 3 dips), 95% ethanol (2 to 3 dips), and tap water. Hematoxylin was applied to the tissue sections for 6 minutes to enhance cellular nuclei, followed by a 2 minute rinse to remove excess stain. A brief 1% Hydrochloric acid solution was applied for 5 seconds in order to enhance staining specificity, followed by a 1 minute wash to remove residual chemicals. Next, a 0.05% ammonia wash was performed for 1 minute, followed by a 1 minute rinse to remove any remaining chemicals. Dehydration of the tissue was accomplished using 95% ethanol for 30 seconds. Eosin was then applied for 3 minutes to stain cytoplasmic structures, followed by an additional tissue dehydration step with 100% reagent alcohol for 3 stations (20 seconds, 30 seconds, 30 seconds). Lastly, the tissue sections underwent clearing with XYLENE for 4 stations, 1 minute each.

#### Bielschowsky silver staining

Slides were deparaffinized and hydrated in distilled water. The Bielschowsky Silver staining was conducted using the Leica ST5010 Autostainer XL. Two solutions of silver nitrate were prepared, a 20% solution (10 g silver nitrate in 50 mL distilled water) and a 10% solution (10 g silver nitrate in 100 mL distilled water). The 20% silver nitrate solution was heated to 60°C for 15 minutes, then the slides were incubated in the heated solution for 15 minutes. Slides were rinsed in distilled water. A formalin solution was prepared of formaldehyde (37-40%) (2 mL) in distilled water (98 mL). A sodium carbonate solution was prepared by mixing 8g sodium carbonate in 30 mL distilled water. **An ammoniacal silver solution was** prepared by treating the 10% silver solution with ammonium hydroxide until the precipitate had almost disappeared, followed by adding sodium carbonate solution (0.5 mL) and ammonium hydroxide (25 drops). The formalin solution was added to the ammoniacal silver solution. This solution was poured over slides and developed for 5 to 30 minutes until golden brown. Slides were rinsed in water and placed in sodium thiosulfate (5 g thiosulfate in 100 mL water) solution for 2 minutes, then washed, dehydrated, cleared, and mounted with synthetic resin.

#### Lewy body scoring (AS)

A laboratory technician blinded to the clinical histories of living participants and postmortem donors reviewed each whole slide image for the presence and density of Lewy bodies. The technician assigned a numerical score to each whole slide image based on the following grading system: 0 (no evidence of Lewy neurites or bodies), 1-(some Lewy neurites, no Lewy bodies), 1 (1 Lewy body), 1+ (2 Lewy bodies), 2- (3-6 Lewy bodies), 2 (7-9 Lewy bodies), 2+ (10-12 Lewy bodies), 3- (13-15 Lewy bodies), 3 (16-18 Lewy bodies), or 3+ (>= 19 Lewy bodies). Scoring criteria were relative to tissue sections of size 1500 μm x 1500 μm and were adjusted for tissue sections of larger or smaller size.

#### Intracellular β-amyloid, cerebral amyloid angiopathy (CAA), and Aβ plaques scoring (4G8)

A laboratory technician blinded to the clinical histories of living participants and postmortem donors analyzed each whole slide image to evaluate the presence and density of intracellular β-amyloid, cerebral amyloid angiopathy (CAA), focal Aβ plaques, and diffuse Aβ plaques. Whole slide images were assigned a final numerical score based on the following grading system: 0 (no evidence of intracellular β-amyloid/angiopathy/Aβ plaques, or white matter only), 1- (intracellular β-amyloid only), 1 (1 instance of angiopathy or plaque, with or without intracellular β-amyloid), 1+ (2-8 instances of angiopathy or plaque, with or without intracellular β-amyloid), 2- (9-15 plaques, with or without intracellular β-amyloid; if there is angiopathy, increase the score value by one), 2 (15-18 plaques, with or without intracellular β-amyloid; if there is angiopathy, increase the score value by one), 2+ (19-25 plaques, with or without intracellular β-amyloid; if there is angiopathy, increase the score value by one), 3- (26-35 plaques, with or without intracellular β-amyloid; if there is angiopathy, increase the score value by one), 3 (>= 36 plaques, with or without intracellular β-amyloid; if there is angiopathy, increase the score value by one), and 3+ (>= 36 plaques and angiopathy, with or without intracellular β-amyloid). Scoring criteria were relative to tissue sections of size 1500 μm x 1500 μm and were adjusted for tissue sections of larger or smaller size.

#### Neurofibrillary tangles (NFTs), glial fibrillary tangles (GFTs), and neuropil thread scoring (AT8)

A laboratory technician blinded to the clinical histories of living participants and postmortem donors reviewed each whole slide image for the presence and density of neurofibrillary tangles (NFTs), glial fibrillary tangles (GFTs), and neuropil threads. Whole slide images were given a final numerical score of 0 (no evidence of NFTs/GFTs/neuropil threads, or white matter only), 1- (neuropil threads only), 1 (1 NFT), 1+ (2 NFTs), 2- (3-6 NFTs, or 1-3 instances of both NFTs and GFTs), 2 (7-9 NFTs, or 4-6 instances of both NFTs and GTFs), 2+ (10-12 NFTs, or 7-9 instances of both NFTs and GFTs), 3- (13-15 NFTs, or 10-12 instances of both NFTs and GFTs), 3 (16-18 NFTs, or 13-15 instances of both NFTs and GFTs), or 3+ (>=19 NFTs, or 16+ instances of both NFTs and GFTs). Scoring criteria were relative to tissue sections of size 1500 μm x 1500 μm and were adjusted for tissue sections of larger or smaller size. Pretangles were not taken into consideration during the scoring process.

#### Gray matter, white matter, and myelin pallor scoring (LHE)

A laboratory technician blinded to the clinical histories of living participants and postmortem donors reviewed each whole slide image for the presence of gray matter, white matter, and myelin pallor. The features were indicated as present or absent using Y/N/NA/X markers. Scoring criteria were relative to tissue sections of size 1500 μm x 1500 μm and were adjusted for tissue sections of larger or smaller size.

#### Neurofibrillary Tangles (NFTs) and Neuritic plaques scoring (Bielschowsky silver)

A laboratory technician blinded to the clinical histories of living participants and postmortem donors reviewed each whole slide image for the presence of Neurofibrillary Tangles (NFTs) and Neuritic plaques. The features were indicated as present or absent using Y/N/NA/X markers. Scoring criteria were relative to tissue sections of size 1500 μm x 1500 μm and were adjusted for tissue sections of larger or smaller size. Non-neuritic amyloid plaques were considered during the scoring process.

### Whole genome sequencing (WGS)

#### DNA extraction and sequencing

DNA was extracted from a total of 500 samples from 500 individuals and all 500 samples were sent for whole genome sequencing after passing quality control assessments. Samples used for DNA extraction were either brain tissue, whole peripheral blood, or fibroblast cell lines. Kits used for extraction included the MagMAX DNA Multi-Sample Ultra Kit 2.0 (ThermoFisher, #A36570), the PAXgene Blood DNA Kit (Qiagen, #761133), and the DNeasy Blood and Tissue Kit (Qiagen, #69504). After extraction, DNA concentration was determined using the Qubit™ dsDNA HS kit (ThermoFisher, #Q32851).

Extracted DNA was sequenced with the Sequencing Core at Vanderbilt University Medical School and whole genome sequencing data was generated for 251 living subjects and 248 postmortem subjects (499 total). Once the genomic DNA arrived at the core facility, a quantitation and integrity assessment were completed. An aliquot of each sample was analyzed on the Agilent TapeStation and quantitated using a Picogreen assay. The samples were normalized and plated using the BioMek FX liquid handler. Whole genomic libraries were prepared using 300 ng of DNA and the Illumina DNA PCR-Free Prep Kit (Illumina, Cat: 20041794) per manufacturer’s instructions. Individual libraries were assessed for quality using the Agilent 2100 Bioanalyzer and quantified with a Qubit Fluorometer. The adapter ligated material was evaluated using qPCR prior to normalization and pooling for sequencing on the QuantStudio 12K Flex. The libraries were sequenced using the NovaSeq 6000 instrument with 150 base-pair paired end reads to a target coverage of 30 reads per base pair. RTA (version 2.4.11; Illumina) was used for base calling.

#### Alignment and variant calling

The Illumina Dynamic Read Analysis for Genomics (DRAGEN) Bio-IT Platform v3.2.8 was used to perform alignment and the first step of variant calling on raw sequencing reads from the DNA WGS samples. Paired-end reads stored in FASTQ files were input into the DRAGEN pipeline. The pipeline was implemented through the “dragen” function in Unix. The “--enable-map-align-output true” argument to the dragen function produced a BAM file for each individual that was sorted by reference sequence and position by default. Duplicate reads were marked in the FLAG field of each BAM file using the “--enable-duplicate-marking true” argument. Read contamination was estimated on the BAM file generated by DRAGEN for each individual using VerifyBamID2 (v1.0.6)^67^.

Haplotype-based variant calling was performed for each individual using the “--enable-variant-caller true” argument, producing a gVCF file for each sample that contained genotype information for all sites in the reference genome detected. Sites where non-reference alleles were detected were considered “variants”. The set of variants generated for each individual were considered the “pre-called” set of variants for the individual.

Genome Analysis Toolkit (GATK; v4.2.0.0) was then used to process pre-called sets of variants in individual gVCFs into a joint-called set of variants. Pre-called sets of variants in individual gVCFs were consolidated into a database using the GenomicsDBImport function of Genome Analysis Toolkit (GATK; v4.2.0.0) and joint genotyping was performed on the database using the GATK GenotypeGVCFs function, producing a single VCF with a record for every site where at least one individual in the cohort was identified as harboring a variant. Sites in the joint-called VCF where multiple alternate alleles were found across the cohort were split into multiple records using bcftools (v1.12).

The GATK Variant Quality Score Recalibration workflow was then implemented (https://gatk.broadinstitute.org/hc/en-us/articles/360035531112--How-to-Filter-variants-either-with-VQSR-or-by-hard-filtering). Variants with a phred-scaled p-value for an exact test of excess heterozygosity (i.e., “ExcessHet” value) greater than 54.69 were removed using the GATK VariantFiltration function. A “sites-only” VCF was created with the GATK MakeSitesOnlyVcf function and VQSLOD values (the log odds of a variant being a true variant versus being a false variant under a trained gaussian mixture model) were analyzed to create VQSLOD tranches (separately for indels and SNPs) using the GATK VariantRecalibrator function. For indels, a “--max-guassians” value of 1 was used for chromosome Y and of 4 was used for all other chromosomes. For SNPs, values of 5 and 6 were used for chromosome Y and all other chromosomes, respectively. Resources used for training the gaussian model were those recommended by GATK Best Practices. The resulting tranches and the sites-only VCF were then processed using the GATK ApplyVQSR function to filter indels and SNPs based on VQSLOD values using the “--truth-sensitivity-filter-level 99.7” argument.

For autosomes, the GATK Genotype Refinement workflow was then implemented (at time of writing, this workflow is not supported for sex chromosomes) (https://gatk.broadinstitute.org/hc/en-us/articles/360035531432). Genotype Quality (GQ) scores were recalculated for each variant called in each individual using the GATK CalculateGenotypePosteriors function. Variants with a recalculated GQ score less than 20 were flagged using the GATK VariantFiltration function, and variants with the resulting flag were set to missing using the GATK SelectVariants function.

#### Data quality control

Following Genotype Refinement, an initial round of variant-level quality control was performed that will be referred to as the “QC0” workflow. The joint-called VCFs from before and after the application of the Genotype Refinement workflow steps were processed using the query function of bcftools to convert the information stored in the INFO fields of the VCFs into tab-delimited files for analysis. Using R (v4.0.3), records that met any of the following criteria were identified for removal: allele count (AC) value of 0 prior to Genotype Refinement; AC value of 0 after to Genotype Refinement; AC value of missing after Genotype Refinement; missing genotypes for greater 5% of the cohort after Genotype Refinement; multiple alternate alleles identified in the cohort; small insertions and deletions. For these records, flags for removal were created in the INFO field of the VCF output of Genotype Refinement using vcfanno (v0.3.3), then the filter function in bcftools was used to update the FILTER field of the VCF based on the newly created flags in the INFO field. The view function in bcftools was then used to only retain records with a “PASS” value in the FILTER field after it had been updated. A total of 32,652,779 records remained in the VCF following implementation of the QC0 workflow.

The VCF output by the QC0 workflow was converted to a plink-formatted binary fileset with plink (v1.90b6.21). Individuals with an estimated contamination greater than 0.02 (N = 49) were removed from the fileset using the “--remove” argument in plink. The plink-formatted QC0 callset was further processed to determine sex from the X chromosome homozygosity estimates. To do this, SNPs were removed for having genotype missingness of greater than 2% and minor allele frequency of less than 5%. Sex was then determined using the plink “--check-sex” argument. Individuals with X chromosome homozygosity estimates less than or equal to 0.25 were considered genetically female. Individuals with X chromosome homozygosity estimates greater than or equal to 0.75 were considered genetically male. Individuals with X chromosome homozygosity estimates greater than greater than 0.25 and less than 0.75 were considered genetically sex ambiguous. Two individuals were identified as “sex mislabels” where the genetically-determined sex was the opposite of the sex recorded in the project database. These two were retained in downstream analyses after correcting the sex mislabels. Two individuals were identified as genetically sex ambiguous and were removed from further analysis.

For each remaining individual, variants were then further subjected to the following filtering steps (“QC1” workflow). For each individual, information about each of the 32,652,779 records in the VCF after applying the QC0 workflow was extracted from the FORMAT field and read into R (v4.0.3). Alternate allele fractions (AAF) were calculated for each record for each individual as the allelic depth (AD) value for the alternate allele divided by the sum of the AD values for the reference and alternate alleles. Records that met any of the following criteria were flagged to have genotypes set to missing for the individual in downstream steps: genotype (GT) calls of 1/0; chromosome Y calls in genetically-determined females (F value<0.25); chromosome X or Y calls in genetically-ambiguous individuals (0.25 < F value < 0.75); for sites expected to be diploid (i.e., autosomes and chromosome X in genetically-determined females) – approximate read depth (DP) value less than 10, homozygous reference genotypes with AAF greater than 0.10, homozygous alternate genotypes with AAF less than 0.90, heterozygous genotypes with AAF less than 0.30 or greater than 0.70; for sites expected to be haploid (i.e., chromosome X and Y in genetically-determined males [F value > 0.75]) – diploid GT calls, approximate read depth (DP) value less than 5, reference genotypes with AAF greater than 0.10, alternate genotypes with AAF less than 0.90

Genotypes flagged to be set to missing by the QC1 workflow were then set to missing using the “--zero-cluster” and “--within” arguments (which provide the list of sites to set genotypes to missing for each individual) in plink. Missingness rates were then calculated for each site in the remaining cohort. At this stage, Y chromosome calls were removed from downstream consideration. Missingness rates for autosomes were calculated in the cohort of 449 individuals remaining at this stage (499 to start minus 49 for contamination and 1 for ambiguous sex). Missingness rates for X chromosomes calls were calculated in the 162 genetically-determined females (though the genotype filters for X chromosomes calls had been applied in males and females) - a step that would allow X chromosome variants to be retained for analysis (otherwise, the DP filter would result in 99% of these calls being removed since at least 2% of males would be marked as missing from low coverage at these haploid sites). Using these calculations of missingness rates, sites with missingness rates greater than 2% were removed from further analysis. Following this filter, all sites with allele count of 0 following these filters were also removed. The resulting set of 25,058,902 sites in 449 individuals is referred to as the “QC1 callset”. Individual-level summary statistics were calculated on the QC1CALLSET using PICARD CollectVariantCallingMetrics and plink –missing, and based on manual evaluation of these metrics one sample was removed for being an outlier with respect to multiple quality control metrics. Identity concordance was performed using all sequencing data performed for the LBP (see identity concordance section below) and 3 additional samples were removed either due to the inability to determine the true identity of the sample (N=2) or for being a duplicate of another sample in the dataset (N=1). The remaining 445 individuals and 25,058,902 sites constituted the “QC2” callset that was the basis for all of the analyses presented here unless additional filters were noted.

### Single-nucleus RNA sequencing (snRNA-seq)

The methods used to perform RNA extraction, RNA sequencing, and data quality control for the LBP snRNA-seq dataset analyzed in the current report are introduced and fully described in Vornholt et al.^17^ and will be briefly summarized here. Single-nucleus suspensions were created using the Minute Single Nucleus Isolation Kit for Neuronal Tissues and Cells (Invent Biotechnologies, #BN-020) following the manufacturer’s protocol. Library construction was performed on single-nucleus suspensions using the Chromium Next GEM Single Cell 3’ Reagent Kits v3.1 (10X Genomics, #CG000204 Rev D) following the manufacturer’s protocol. The libraries were pooled for sequencing and checked for RNA quality and RNA quantity with the Agilent TapeStation. Sequencing was done on the Illumina NovaSeq using the NovaSeq 6000 S2 Reagent Kit v1.5 kit (Illumina, #20028315) to a target of 300 million reads per sample. Raw FASTQ files were generated from successfully sequenced libraries. The CellRanger software (pipeline version 7.0) was used for alignment of reads to the reference transcriptome (version GRCh38-2020-A), generation of raw unique molecular identifier (UMI) counts, and nuclei filtering. The number of nuclei retained for each sample after the filter applied by CellRanger was calculated, and the distribution of these values was manually assessed to identify samples that were outliers to be excluded from downstream analyses. Removal of contaminating ambient RNA was done via SoupX, which estimates the cell-free expression profile and background-corrects unique molecular identifier (UMI) count matrices accordingly. Empty droplets and low-quality nuclei were removed from background-corrected count matrices via Seurat (version 3.0) if (1) total genes and counts were greater than 200 or (2) more than 1% of counts originated from mitochondrial RNA. Doublets were identified and removed using scDblFinder (ver. 3.16). The output of these quality control procedures was 362,310 nuclei from 31 LIV samples and 21 PM samples.

Background-corrected UMI matrices from the samples that remained after sample filtering were merged together into a single matrix including 362,310 nuclei from 52 samples (31 LIV samples and 21 PM samples). These counts were normalized to counts-per-million (CPM) using the RelativeCounts() function of the Seurat R package with a scale factor of 10^6^. A value of 1 was added to each value in the CPM matrix, and these values were then log2-normalized using the log2() function of the base stats R package. The 2,000 most highly variable genes were identified in the log2-normalized matrix using the FindVariableFeatures() function of the Seurat R package with the “selection.method” parameter set to “vst.” The log2-normalized counts for these 2,000 genes were then adjusted to account for the unwanted effects of technical variables (i.e., number of counts, number of features, percentage of counts mapping to exons) on gene expression using the ScaleData() function of the Seurat R package. Principal components analysis was performed on the adjusted log2-normalized counts using the RunPCA() function of the Seurat R package. The effects of sample identity on the top 15 principal components was accounted for by adjusting the top 15 principal components using the RunHarmony() function of the harmony R package (version 1.1). The harmony-adjusted principal components were then input into the FindNeighbors() and FindClusters() functions of the Seurat R package to identify clusters of nuclei, with the “resolution” parameter of the FindClusters() function set to 0.6.

Cell clusters were then annotated into 6 broad classes: excitatory neurons [Exc]; inhibitory neurons [Inh]; oligodendrocytes [Oli]; astrocytes [Ast]; microglia [Micro]; oligodendrocyte progenitor cells [OPCs]. Annotations were made by comparing clusters to clusters of the same nuclei previously annotated in the Vornholt et al report^17^. The clustering solution for this report differs slightly from the clustering solution in Vornholt *et al.* because in this report the effect of technical variables were regressed out prior to clustering.

Differential expression analyses performed using the snRNA-seq data utilized “pseudo-bulk” counts. For each cell type studied (i.e., Exc, Inh, Oli, Ast, Micro, OPC), a pseudo-bulk count matrix was calculated using aggregateData() function of the muscat R package (v1.10.1) that had one column for every sample and one row for every gene detected in the nuclei annotated to that cell type. The values in these matrices were the sum of the background-corrected UMI counts for the gene in the nuclei from the sample annotated to the cell type.

### Identity concordance

To identify sample mislabeling events, identity concordance between samples was performed using genetic variants called from the bulk RNA-seq data, WGS data, snRNA-seq data, and RNA sequencing data from blood samples from the same individuals generated for other ongoing LBP studies. More than one data source was available for identity concordance for all but one individual. Variants were called from RNA sequencing data following Genome Analysis Toolkit (GATK) best practices. The approach used to determine if two variant call sets were from the same individual differed depending on the sources of the call sets in the comparison (e.g., RNA sequencing data to RNA sequencing data, WGS to RNA sequencing data). For comparisons of (1) two call sets from bulk RNA sequencing data, (2) two callsets from single-cell RNA sequencing data, and (3) two call sets from WGS data, gtcheck from the bcftools software package (v1.9) was used to calculate the percentage of sites concordant between the call sets. For comparisons of (1) bulk RNA sequencing data call sets to single-cell RNA sequencing data call sets, (2) WGS data callsets to bulk RNA sequencing data call sets, and (3) WGS data callsets to single-cell RNA sequencing data call sets, genotyping matrices were read into R, discrepancies between allele code fields were corrected, sites covered in only one call set were removed, and Pearson’s correlations of genotype similarity were calculated for every pair of call sets. Regardless of the approach used to calculate concordance between call sets, a threshold for deciding whether two samples came from the same individual was determined manually by assessing the distribution of similarity metrics. Mismatches were defined as (1) instances where two samples expected to be from the same individual were genetically discordant and (2) instances where two samples not expected to be from the same individual were genetically concordant. All mismatches identified by the thresholding procedure were further examined to confirm a true mismatch (i.e., using all RNA sequencing data and WGS data call sets from the individuals in the mismatch).

### FSCV DE analyses

Each of the pseudo-bulk count matrices was filtered to retain the 15 LIV samples with FSCV recordings. Lowly expressed genes were then removed from each matrix using the default settings of the filterByExpr() function in the edgeR R package (v3.38.1). The pseudo-bulk counts of remaining genes were normalized using the voomWithDreamWeights() function of the dream software within the variancePartition R package. A total of 18 FSCV DE analyses were performed (one in each of the six cell types for each of the three neurotransmitters [dopamine, serotonin, norepinephrine]). For each of these 18 FSCV DE analyses, DE was run on the voom-normalized pseudo-bulk count matrix using the dream() function in the variancePartition R package and the formula indicated below. For all FSCV DE analyses, multiple test correction was done using an FDR of 5%. FSCV DEFs were defined using an unadjusted p-value threshold of 0.05. FSCV DE signature π_1_ statistics^32^ were estimated using the qvalue R package (v2.28.0). KEGG enrichment of FSCV DEFs was performed as described below.

<u>Formula 2 (linear model with fixed effects and random effects for individual ID and and sex):</u>

*RNA Transcript Expression ∼ FSCV Metric + Individual ID + Sex + Number of Nuclei + Mean Reads Per Nucleus + Fraction of Reads in Nuclei + Residuals*

### KEGG enrichment tests

Gene set enrichment analyses were performed using the Kyoto Encyclopedia of Genes and Genomes (KEGG) database^33^. The htext-formatted hsa00001 KEGG database was downloaded on October 4th, 2021, via the KEGG web server and parsed into structured data tables in python. The output of this script was a row for every instance of a feature’s membership in a KEGG gene set and one column each for gene symbol, gene set (“KO reference pathway”), parent category of the gene set (“super pathway”), and the broad concept grouping of the parent category (“top-level string”). Gene symbols provided by KEGG were mapped to Ensembl identifiers with a mapping file generated using the HUGO^68^ web server on June 10th, 2020. All KEGG gene sets with the top-level string “Human” were excluded from analysis since these gene sets may be derived from studies of postmortem human tissues. Sets with less than 10 member genes were excluded from analysis, leaving 278 KEGG gene sets tested for enrichment. These 278 KEGG gene sets were tested for enrichment of DEFs in R using the fisher.test() function in the base stats package as the overlap between the genes in the KEGG gene set and DEFs, using as background the set of features (i.e., RNA transcripts or proteins) in the DE analysis. KEGG gene sets with statistically significant associations were mapped to parent terms via the source data file. Multiple testing correction was carried out separately for each of the DE signatures investigated accounting for the 278 KEGG gene sets tested using the false discovery rate estimation method of Benjamini and Hochberg^69^ implemented in R using the p.adjust() function of the base stats package.

### Comparing DE signatures

Spearman’s correlations between pairs of DE signatures presented throughout this report were calculated using the cor.test() function in the R base stats package. Multiple test correction for these Spearman’s correlations was not performed because the majority of unadjusted p-values returned by the cor.test() function were estimated to be 0 (indicated in the main text using the “p-value < 2.2 x 10^−16^” notation). When throughout this report the overlap between two DEF sets was compared using Fisher’s exact test (e.g., FSCV DEFs and iEEG DEFs), the Fisher’s exact test was implemented by the fisher.test() function in the base stats R package.

### MER DE analyses

DE of the aperiodic exponent values was performed on the voom-normalized mature RNA transcript bulk RNA-seq data separately for the STN (N = 72 LIV samples from 54 individuals) and GPi (43 LIV samples from 28 individuals) using the dream() function in the variancePartition R package and the formula indicated below. The covariates to include in the formula were selected based on the covariate selection process performed for the LIV-PM bulk RNA-seq DE analyses with the exception that for MER DE the estimated percentage of neuronal cells was not used as a covariate in order to maximize the comparability between the MER DE and the snRNA-seq FSCV DE signatures. For all MER DE analyses, multiple test correction was done using an FDR of 5%. MER DEFs were defined using an unadjusted p-value threshold of 0.05. MER DE signature π_1_ statistics^32^ were estimated using the qvalue R package (v2.28.0).

<u>Formula 3 (linear model with fixed effects and random effects for individual ID and and sex):</u>

*RNA Transcript Expression ∼ Aperiodic Exponent + Individual ID + Sex + RIN + Picard RNASeqMetrics Median 3 Prime Bias + Picard RNASeqMetrics PCT mRNA Bases + Depletion Batch + Picard InsertSizeMetrics Median Insert Size + Alignment Summary Metrics + Residuals*

### Intracranial electroencephalography (iEEG) DE analyses

RNA transcript associations with neural oscillations measured using iEEG were obtained from Supplementary Table 2 of the Berto *et al.* publication (sheet entitled “SME Univariate Stats”)^19^. Gene symbols were mapped to ensembl gene identifiers using HUGO. Since the published associations were already limited to genes with significant associations (i.e., IEEG DEFs), no further filtering was done (2,380 unique ensembl gene identifiers).

### Identification of TPAWN

To identify the set of RNA transcripts referred to as “TPAWN” in the results, the following procedure was performed. The initial input to the procedure was 18 neurotransmission DE signatures (the 10 FSCV DE signatures with π_1_ values greater than 0.10; the two MER DE signatures; and the 6 iEEG DE signatures). Two DEF lists were made from each of the DE signatures: Up DEFs (i.e., DEFs with positive logFC values) and Down DEFs (i.e., DEFs with negative logFC values). As described above, for the FSCV and the MER DE signatures DEFs were defined using unadjusted p-value thresholds and for the IEEG DE signatures DEFs were defined in the Berto *et al.* publication^19^. Each of the 18 neurotransmission DE signatures was compared to the other 17 neurotransmission DE signatures by performing the following 4 Fisher’s exact tests (for a total of 612 tests):

1. Up DEFs from the first DE signature were compared to Up DEFs in the second signature
2. Up DEFs from the first DE signature were compared to Down DEFs in the second signature
3. Down DEFs from the first DE signature were compared to Up DEFs in the second signature
4. Down DEFs from the first DE signature were compared to Down DEFs in the second signature

For comparisons of two LBP neurotransmission DE signatures, the set of background features used in the Fisher’s exact test was the intersect between the features present in the two DE signatures. For comparisons between a LBP neurotransmission signature and a IEEG DE signature, the set of background features used in the Fisher’s exact test was the set of features present in the LBP DE signature. This was necessary because the full DE signature (i.e., logFCs from all features tested) was only available for LBP neurotransmission DE signatures (i.e., the full IEEG DE signatures were not published, only the iEEG DEFs were published). For comparisons between two IEEG DE signatures, the set of 30,099 RNA transcripts expressed in the bulk RNA-seq mature RNA DE analyses were used as the background. After correcting for 612 tests using the FDR of 5%, any two signatures with at least one significant test with an odds ratio greater than 1 were considered “connected” to one another, resulting in 55 connections between 17 neurotransmission DE signatures. These connections served as the input to construct an undirected graph using the igraph R package (v1.4.1), where nodes were the 17 DE signatures and edges were the 55 connections between these signatures. Fully connected subcomponents of the graph were identified using the maximal.cliques() function of the igraph R package, and the resulting list of subcomponents was filtered to include only those subcomponents that included a node from each of the three types of neurotransmission DE signatures (i.e., FSCV, MER, and iEEG). Only one such subcomponent was identified. The union of the DEFs from the nodes in this subcomponent was then compiled, and features that were DEFs in signatures in at least two types of neurotransmission DE (i.e., FSCV and MER; FSCV and iEEG; MER and iEEG) were considered part of the TPAWN (N = 588 RNA transcripts).

### Enrichment of TPAWN genes in relevant external data

#### NeuroExpresso/NeuroElectro Data

The NeuroExpresso/NeuroElectro study^36^ associated RNA transcripts to neurotransmission by integrating RNA transcript expression data generated using microarray technology from mouse brain tissue with data manually curated from the literature characterizing electrophysiological properties of rodent brain cells. A set of approximately 1,000 RNA transcripts identified by the NeuroExpresso/NeuroElectro study as being significantly associated with an electrophysiological property was downloaded (i.e., the list of RNA transcripts the NeuroExpresso/NeuroElectro study team published in Supplementary Table 3 of the NeuroExpresso/NeuroElectro study report^36^). A Fisher’s exact test was performed using the fisher.test() function in the base stats R package to assess the overlap between these RNA transcripts and the RNA transcripts in TPAWN. The background set of genes used in the Fisher’s exact test was the 30,099 genes expressed in the PFC in the bulk RNA-seq mature RNA transcript expression data.

#### Allen Institute for Brain Science Data

Published analyses of data from the Allen Institute for Brain Science^37^ associating RNA transcript expression to electrophysiological properties were downloaded (“Online Table 1” at https://github.com/PavlidisLab/transcriptomic_correlates). The downloaded table contained all associations between a RNA transcript and a electrophysiological property identified by the authors. The overlap between the set of associated RNA transcripts in the downloaded table and the RNA transcripts in TPAWN was assessed using a Fisher’s exact test was performed using the fisher.test() function in the base stats R package. The background set of genes used in the Fisher’s exact test was the 30,099 genes expressed in the PFC in the bulk RNA-seq mature RNA transcript expression data.

#### Evolutionary constraint

Evolutionary constraint of genes was defined using “loss-of-function observed/expected upper bound fraction” (LOEUF) scores from the Genome Aggregation Database (gnomAD)^35^. The distribution of LOEUF scores in genes encoding the 588 RNA transcripts in TPAWN was compared to distribution of LOEUF scores in all other genes expressed in Exc of the snRNA-seq data using the ks.test() function in the base stats R package.

#### GWAS catalog

Associations between genes and traits were downloaded from the GWAS catalog (v1.0.2)^70^ on December 12^th^, 2023. For each trait, a Fisher’s exact test was performed using the fisher.test() function in the base stats R package to assess the overlap between the genes associated with the trait through GWAS and the genes encoding the RNA transcripts in TPAWN. Multiple test correction for the number of traits in the catalog was performed using the method of Benjamini and Hochberg implemented in R using the p.adjust() function of the base stats package. Only traits with at least 10 genes associated through GWAS were retained for analyses. The background set of genes used in the Fisher’s exact test was the 30,099 genes expressed in the PFC in the bulk RNA-seq mature RNA transcript expression data.

#### Mental health phenome-wide association study

Genetic and clinical data from the Mount Sinai Million Health Discoveries Program (MSM-HDP; formerly, the BioMe Biobank Program; N = 29,064) were analyzed to test the effects of rare PTVs in the genes encoding RNA transcripts in TPAWN on illnesses of higher-order brain functions. The genetic data was comprised of whole-exome sequencing (for identifying rare PTVs) and microarray (for calculating genetic principal components); the clinical data was comprised of billing codes from the electronic medical records that had been mapped to phecodes^71–74^. Variants in the MSM-HDP call set were annotated for their predicted effect on protein function using the Variant Effect Predictor (VEP; v96) software with the LOFTEE plugin. PTVs were defined as any variant annotated by the LOFTEE plugin as “loss-of-function” with either “high confidence” or “low confidence.” PTVs were defined as rare if the minor allele count in the MSM-HDP cohort was less than or equal to five. A phenome-wide association study (PheWAS) was run using the phewas() function of the PheWAS() R package (v0.99.5-5) to test the association between each phecode and being a carrier of a rare PTV in one of the genes encoding RNA transcripts in TPAWN. The top 20 genetic principal components were used as covariates. After running the PheWAS, phecodes were grouped into broad categories using the phewasManhattan() function of the PheWAS R package, and the 68 phecodes in the “mental disorders” category were retained for analysis. Multiple test correction for these 68 phecodes was performed using the method of Benjamini and Hochberg implemented in R using the p.adjust() function of the base stats package.

### Annotating TPAWN in Exc to Npr3 Exc neurons

A score representing the activity of the 588 RNA transcripts in TPAWN in each nucleus of the snRNA-seq data from LIV samples (N = 31; N = 60,834 nuclei) was calculated using the AUCell_run() function of the AUCell R package (v1.18.1). For visualization of these 60,834 nuclei in UMAP dimensions, the same procedure was followed that was described above for the full set of 362,310 nuclei. The mean TPAWN score amongst these 60,834 nuclei was calculated. Nuclei with scores greater than the mean were labeled as having high TPAWN scores and nuclei with scores lower than the mean were labeled as having low TPAWN scores. The 60,834 Exc nuclei were annotated with respect to transcriptional signature of mouse PFC excitatory neurons with known axonal projection patterns. These signatures were downloaded from a published study^38^, and a score for each signature was calculated using the AUCell_run() function of the AUCell R package (v1.18.1). The relationship between the mouse PFC projection neuron scores and the TPAWN scores was assessed using linear regression implemented with the lm() function of the base stats R package. The TPAWN score was the dependent variable in the linear regression model and the mouse PFC projection neuron scores were the dependent variables. To visualize TPAWN scores against Npr3 scores, a score for the Npr3 signature was re-calculated using the AUCell_run() function of the AUCell R package (v1.18.1) after removing features from the Npr3 signature that were also in TPAWN.

### LIV-PM DE in sn-RNAseq data

LIV-PM DE in the snRNA-seq data was performed for each cell type in order to compare FSCV DE signatures to a LIV-PM DE signature of the same cell type from an independent set of samples. Three filters were applied to each pseudo-bulk count matrix (31 LIV samples and 21 PM samples) to create the datasets for LIV-PM DE: (1) the 15 LIV samples in the FSCV DE analyses were removed; (2) one of the PM samples that was a technical replicate of another PM sample was removed; (3) for living participants with two LIV samples, one sample was removed. These filters resulted in 13 LIV samples and 20 PM samples (one sample per individual) for analysis. Lowly expressed genes were then removed from the pseudo-bulk counts matrices using the default settings of the filterByExpr() function in the edgeR R package (v3.38.1). Since these matrices consisted of one sample per individual, the pseudo-bulk counts of expressed genes were normalized using the voom() function of the limma R package (v3.46.0)^75^ and DE of LIV-PM status was performed using the lmFit() function of the limma R package using the following formula (determined as described in Vornholt *et al.*^17^):

<u>Formula 4 (linear model with fixed effects and random effects for individual ID and sex):</u>

*RNA Transcript Expression ∼ LIV-PM Status + Individual ID + Sex + Number of Nuclei + Mean Reads Per Nucleus + Fraction of Reads in Nuclei + Residuals*

### Calculating residual bulk RNA-seq and bulk LC-MS expression levels

Residual bulk primary RNA transcript, mature RNA transcript, and protein expression data was calculated using the following formulas applied to the normalized counts using the residuals() function of the base stats R package:

<u>Formula 5 (linear model with fixed effects and random effects for individual ID, sex, and depletion batch):</u>

*RNA Expression ∼ Individual ID + Sex + RIN + Neuronal Fraction + Picard RNASeqMetrics Median 3’ Bias + Picard RNASeqMetrics PCT mRNA Bases + Depletion Batch + Picard InsertSizeMetrics Median Insert Size + Picard AlignmentSummaryMetrics Strand Balance (First of Pair) + Residuals*

<u>Formula 6 (linear model with random effects for individual ID):</u>

*Protein Expression ∼ Individual ID + Residuals*

After applying these formulas, dimensionality reduction was performed on residual primary RNA, mature RNA, and protein expression data using the Uniform Manifold Approximation and Projection (UMAP) method implemented through the umap() function in the umap R package (v0.2.8.0) with the “n_components” parameter set to 2.

For RNA-protein co-expression analyses, the formula was used to calculate residuals for RNA did not include neuronal cell fraction. As described in the methods section, for both RNA and protein expression data, the process for selecting covariates to include in models consisted of evaluating the effects of each potential covariate on the top PCs of expression data. This process resulted in the neuronal cell fraction estimate being selected for inclusion of RNA data. The neuronal cell fraction is estimated using cell-type-specific references of postmortem human brain gene expression. Such references do not exist for human brain proteomics data. Neuronal cell fraction estimates derived from RNA transcript expression could not be used to estimate neuronal cell fractions from protein expression of the same sample for two reasons: (1) different portions of the sample were used for RNA sequencing and protein, such that the cell type fraction of the portion of the sample used for RNA sequencing may not be representative of the cell type fraction of the portion of the samples used for LC-MS, and (2) the low same-gene correlations between RNA transcript and protein expression from previous studies^11^. Since neuronal cell fraction could only be estimated for one of the data modalities, when performing the analyses integrating the two analyses a decision had to be made regarding whether to include neuronal cell fraction as a covariate in the RNA sequencing data or not. The goal was to make this decision based on what would result in a true representation of the RNA transcript and protein co-expression in the samples, and it was therefore decided to allow cell type-specific effects on expression to remain in the RNA transcript data since these effects could not be accounted for in the protein expression data. Residuals of RNA transcript data for the RNA-protein co-expression analyses were therefore calculated using the following formula:

<u>Formula 7 (linear model with fixed effects and random effects for individual ID, sex, and depletion batch):</u>

*RNA Expression ∼ Individual ID + Sex + RIN + Picard RNASeqMetrics Median 3’ Bias + Picard RNASeqMetrics PCT mRNA Bases + Depletion Batch + Picard InsertSizeMetrics Median Insert Size + Picard AlignmentSummaryMetrics Strand Balance (First of Pair) + Residuals*

### LIV-PM DE on bulk RNA-seq and bulk LC-MS data

For each of the LIV-PM DE analyses performed on the bulk RNA-seq and bulk LC-MS data, DE was run on a normalized count matrix using the dream() function in the variancePartition R package and one of the formulas indicated below. For each DE analysis, multiple test correction was done using an FDR of 5%. The LIV-PM DE analyses performed on the bulk RNA-seq and bulk LC-MS data in this report were as follows:

1. Primary RNA LIV-PM DE [Formula 8, DE Trait: LIV-PM status]
2. Mature RNA LIV-PM DE [Formula 8, DE Trait: LIV-PM status]
3. Protein LIV-PM DE [Formula 9, DE Trait: LIV-PM status]

<u>Formula 8 (linear model with fixed effects and random effects for individual ID, depletion batch, and sex):</u>

*RNA Transcript Expression ∼ DE Trait + Individual ID + Sex + Percentage Neuronal Fraction + RIN + Picard RNASeqMetrics Median 3 Prime Bias + Picard RNASeqMetrics PCT mRNA Bases + Depletion Batch + Picard InsertSizeMetrics Median Insert Size + Alignment Summary Metrics + Residuals*

<u>Formula 9 (linear model with fixed effects and random effects for individual ID):</u>

*Protein Expression ∼ DE Trait + Individual ID + Residuals*

### LIV-PM differential splicing rate analysis on bulk RNA-seq data

Differential RNA splicing rates were tested between LIV samples and PM samples by adapting the procedures used for differential expression of RNA transcript expression levels described above. The starting point for this analysis was the transcript-level counts matrix for primary RNA and mature RNA output by salmon prior to normalization. Only RNA features present in both the primary RNA and mature RNA expression matrices were included (20,671 RNA transcripts).

In order to normalize the primary RNA and mature RNA matrices together, after subsetting the primary RNA and mature RNA matrices for these features, the two matrices were combined in two different ways to construct a “long” form and a “wide” form. The long form was constructed by concatenating the primary RNA matrix (20,671 rows by 518 columns) and the mature RNA matrix (20,671 rows by 518 columns) into a single matrix of 41,342 rows and 518 columns. In this form, there were two rows for each RNA transcript, and the row name for these two rows were differentiated from each other using an indicator of whether the row represented the primary RNA or the mature RNA for of the transcript (e.g., “FEATURE1-Primary”, “FEATURE1-Mature”). The wide form was constructed by joining the primary RNA matrix and the mature RNA matrix by RNA transcript name into a single matrix of 20,671 rows and 1036 columns. In this form, there was only one row for each RNA transcript (e.g., “FEATURE1”) but there were two columns for each sample, and the column names for these two columns differentiated from each other using an indicator of whether the sample was from the primary RNA or the mature RNA data (“SAMPLE1-Primary”, “SAMPLE1-Mature”). Normalization factors for each of the 518 samples were calculated on the long form of the matrix using the calcNormFactors() functions of the edgeR R package (v3.38.1). The wide form of the matrix was then normalized using these normalization factors with the voomWithDreamWeights() function of the variancePartition R package.

Differential expression was performed on the wide form of the matrix. To do this, a metadata table needed to be constructed that had one row for every column in the wide form of the expression matrix. The columns in the metadata table included LIV-PM status, technical covariates used in differential expression analyses, and a column that indicated whether the row was from the primary RNA or mature RNA data. The formula (“Formula 10” below) for differential splicing rates included an interaction term between the LIV-PM status of the sample and primary-mature RNA status of the sample. Conceptually, the resulting summary statistics from the dream() function of the variancePartition R package represent the differences in the ratio of mature RNA levels (i.e., spliced RNA) to primary RNA levels (i.e., unspliced RNA) between LIV samples and PM samples for each RNA transcript. Positive logFC values mean PM samples have higher splicing rates than LIV samples, and negative logFC values mean that LIV samples have higher splicing rates than PM samples.

<u>Formula 10 (linear mixed model with fixed effects and random effects for individual ID, depletion batch, and sex):</u>

*Splicing Rate ∼ Spliced Status*LIV-PM Status + Sex + RIN + Neuronal Fraction + Picard RNASeqMetrics Median 3’ Bias + Picard RNASeqMetrics PCT mRNA Bases + Individual ID + Depletion Batch + Picard InsertSizeMetrics Median Insert Size + Picard AlignmentSummaryMetrics Strand Balance (First of Pair) + Residuals*

### LIV-PM differential intron usage analysis on bulk RNA-seq data

Leafcutter (v0.2.9) was used to perform differential intron usage analysis on 12,438 intron clusters that remained following filtering procedures described above. LIV samples for this analysis were downsampled to retain one sample per individual due to the inability of the current version of Leafcutter to model the effect of multiple samples per individual (169 LIV samples and 243 PM samples). The sample to retain was chosen randomly for each individual. Differential intron usage was run using the differential_splicing() function of the leafcutter R package (v0.2.9)^41^ with the “min_samples_per_intron” and “min_samples_per_group” parameters both set to 10 and “confounders” set to a matrix of covariates that included the same covariates used in Formula 1 above with the exception of Individual ID since there was only one sample per individual used here. Of the 12,438 intron clusters input into the differential intron usage analysis, differential intron usage was successfully run on 11,600 (containing 29,002 of the 32,632 introns) – 808 intron cluster tests failed for timing out and 30 failed for not having enough samples meeting the criteria for testing. After mapping the gene identifiers output by Leafcutter (i.e., gene symbols) to ensembl IDs using the Gencode v30 reference transcriptome source files, 11,222 of the 11,600 intron clusters successfully tested were mapped to an Ensembl gene identifier (6,797 unique Ensembl gene identifiers, meaning some Ensembl gene identifiers contained greater than one intron cluster). The 378 intron clusters without an Ensembl gene identifiers also did not have an associated gene symbol, and these were removed from analysis. For visualization of differential intron usage in *RSRP1*, the following procedures were followed. The gtf2leafcutter.pl script of the leafcutter package (v0.2.9) was used to generate files for annotating genes in the format required by subsequent leafcutter scripts, using as input the GTF file from the Gencode v30 release. Next, using the resulting annotation files, the intron counts and differential intron usage summary statistics were annotated to genes using the prepare_results.R script of the leafcutter package (v0.2.9), with the “-f” parameter to define significance of intron usage tests set to 0.05. Using the intron counts matrix returned by the prepare_results.R script, for each intron in the *RSRP1* intron cluster the LIV mean count was calculated as the mean of the counts in LIV samples, and the LIV intron usage was calculated as the LIV mean count for the intron divided by the sum of the LIV mean counts for all introns in the cluster. The same procedure was repeated to calculate the PM intron usage for each intron in the cluster. For introns where the LIV mean count was greater than the PM mean count, the intron usage ratio was calculated as the LIV mean count divided by the PM mean count. For introns where the PM mean count was greater than the LIV mean count, the intron usage ratio was calculated as the PM mean count divided by the LIV mean count.

### RNA-protein correlation difference matrices

For the residual mature RNA (30,099) and protein (6,415) expression data, the following procedure was performed to determine if the feature-feature correlations differed between LIV (N=235 in both RNA and protein) and PM (N=237 in both RNA and protein) samples:

1. Feature-feature Pearson’s correlation matrices were calculated separately for LIV samples and PM samples using the rcorr() function in the Hmisc (v4.7-0) package in R.
2. The LIV sample feature-feature correlations were subtracted from the PM sample feature-feature correlations and transformed to absolute values, resulting in the “LIV-PM correlation difference matrix.”
3. The mean of the LIV-PM correlation difference matrix was computed.
4. An empirical p-value was calculated to assess the significance of the LIV-PM correlation difference matrix mean. The null distribution used to calculate this p-value was generated through 10,000 permutations of the following five-step procedure: 1) LIV samples were randomly split in half and PM samples were randomly split in half; 2) each of the LIV sample halves was combined with one of the PM sample halves, yielding two “random” sets of samples; 3) feature-feature Pearson’s correlation matrices were calculated for each of the random sample sets using the cor() function in the base stats R package (rcorr was not used here because p-values were not needed); 4) the “null correlation difference matrix” was calculated by subtracting the correlation matrix from one random set from the correlation matrix of the other random set and transforming to absolute values; 5) the null correlation difference matrix mean was computed. The empirical p-value was calculated as the fraction of 10,000 null correlation difference matrix means that were greater than the LIV-PM correlation difference matrix mean.

### LIV-PM differential co-expression analyses on bulk RNA-seq and bulk LC-MS data

A mature RNA-protein Pearson’s correlation matrix (30,099 x 6,415) was calculated separately for LIV (N=235 with both RNA and protein) and PM (N=237 with both RNA and protein) samples using the rcorr() function in the Hmisc (v4.7-0) package in R (v4.2.0). A matrix was generated for correlation coefficients and the corresponding p-values. The p-values were adjusted by applying the FDR through the p.adjust() function of the base stats R package (adjusting for 30099 x 6415 = 193,085,085 tests). Adjustment of p-values was performed separately for LIV and PM samples.

RNA-protein pairs were considered “same-gene” pairs if the mature RNA and protein features were derived from the same gene. A total of 5,714 same-gene pairs were present after mapping the 6,415 refseq protein identifiers to ensembl identifiers (successful mapping for 5,758) and subsetting for the ensembl identifiers also present in the 30,099 mature RNA features. The full RNA-protein matrix (30099 x 6415) was subset for these proteins (30099 x 5714 = 171,985,686 RNA-protein correlations).

To test whether same-gene correlations in LIV and PM samples were enriched for being positive and significant, each mature RNA-protein pair was labeled as being 1) a “same-gene” pair and 2) a pair with a statistically significant and positive Pearson’s correlation coefficient (statistical significance here was defined based on the adjusted p-values calculated on the full mature RNA-protein matrix prior to subsetting proteins for the 5714); then, a one-sided Fisher’s exact test was performed to test whether same-gene pairs were enriched for positive-significant pairs using the fisher.test() function in the based stats R package.

The concordance of the same-gene mature RNA-protein correlations in LIV and PM samples was calculated using the cor() function of the base stats R package (Pearson’s). To determine if the LIV-PM status was driving same-gene mature RNA-protein correlations, t-statistics and associated p-values were obtained using the following linear model run using the lm() function of the base stats R package:

<u>Formula 11 (linear model):</u>

*Same-gene correlation coefficient ∼ Mature RNA LIV-PM logFC + Protein LIV-PM logFC*

The resulting p-values (4 p-values total, two from the LIV model and 2 from the PM model, one for each logFC variable) were adjusted using the Bonferroni method in the p.adjust() function in the base stats R package. For plotting these results, the mature RNA logFC and the protein logFC were summed, and if the sum was greater than 0 the point was colored blue and if the sum was less than 0 the point was colored pink, then the shade of the point was set to reflect the absolute value of the sum.

To assess whether mature RNA-protein pair correlations differed between LIV (N=235 in both RNA and protein) and PM (N=237 in both RNA and protein) samples:

1. The LIV sample mature RNA-protein correlations were subtracted from the PM sample mature RNA-protein correlations and transformed to absolute values, resulting in the “LIV-PM mature RNA-protein correlation difference matrix.”
2. An empirical p-value was calculated for each mature RNA-protein pair in the LIV-PM mature RNA-protein correlation difference matrix to assess the significance of the correlation difference. The null distribution used to calculate this p-value was generated through 10,000 permutations of the following four-step procedure: 1) LIV samples were randomly split in half and PM samples were randomly split in half; 2) each of the LIV sample halves was combined with one of the PM sample halves, yielding two “random” sets of samples; 3) mature RNA-protein Pearson’s correlation for the mature RNA-protein pair of interest was calculated for each of the random sample sets using the cor() function in the base stats R package (R version v4.2.0); 4) the “null correlation difference” was calculated by subtracting the correlation from one random set from the correlation of the other random set and transforming the difference to an absolute value. The empirical p-value was calculated as the fraction of 10,000 null correlation differences that were greater than the LIV-PM correlation difference for the pair of interest.

The percentage of mature RNA-protein pairs that were significantly correlated in either LIV samples or PM samples that were also significantly differentially correlated between LIV samples and PM samples was calculated using the adjusted p-values to define pairs significantly correlated in either LIV or PM (adjusted p-values < 0.05) and using the empirical p-values to define the pairs where the LIV correlation was significantly different than the PM correlation (empirical p-value < 0.05). The OR to test whether the mature RNA-protein pairs that were significantly correlated in either LIV samples or PM samples were enriched for being significantly differentially correlated between LIV samples and PM samples was calculated using the fisher.test() function in the base stats R package as follows:

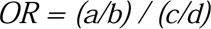

*where,*

*a = Number of pairs significantly correlated in LIV or PM and significantly differentially correlated between LIV and PM (N=1608593)*

*b = Number of pairs significantly correlated in LIV or PM and not significantly differentially correlated between LIV and PM (N= 600205)*

*c = Number of pairs not significantly correlated in LIV or PM and significantly differentially correlated between LIV and PM (N=14783192)*

*d = Number of pairs not significantly correlated in LIV or PM and not significantly differentially correlated between LIV and PM (N=176093095)*

To test if mature RNA-protein correlation differences between LIV and PM samples were enriched for protein families defined in KEGG the following approach was taken. KEGG sets labeled as “Protein Families” in the SuperPathwayString field of the KEGG source files were considered for testing, and after filtering for sets that contained at least 10 proteins in the LBP data 36 sets remained for testing. To create the background for these tests a table was created with one row for each of the 2,208,798 mature RNA-protein pairs that were significantly correlated in either LIV or PM samples, with one column for the RNA feature in the pair and one column for the protein feature in the pair. A column was created indicating whether the mature RNA-protein pair was significantly differently correlated between LIV samples and PM samples. For each of the 36 KEGG sets, a column was created indicating whether the protein feature in the pair was a member of the KEGG set. Finally, a Fisher’s exact test was performed to test whether the rows with a TRUE value in the column for differential correlations were enriched for the rows with TRUE values in the KEGG column. Multiple testing correction was done for the 36 tests using the FDR method in the p.adjust() function in the base stats R package.

### WGS data processing for *cis*-eQTL analyses

For *cis*-eQTL analyses, the additional procedures were performed following the WGS data processing procedures described above.

#### Principal components analysis (PCA)

PCA was performed on WGS data in order to account for ancestry in all *cis*-eQTL analyses. Prior to PCA, the QC2 callset was filtered to remove sites with missingness greater than 2%, minor allele counts less than 10, and Hardy-Weinberg p-value < 5 x 10^−16^. Variants were then further restricted to autosomal sites. Regions of the genome known to be under recent selection were removed: specifically HLA (chr6:27000000-35000000, hg19), LCT (chr2:135000000-137000000), an inversion on chromosome 8 (chr8:6000000-16000000), a region of extended LD on chromosome 17 (chr17:40000000-45000000), EDAR (chr2:109000000-110000000), SLC2A5 (chr15:48000000-49000000), and TRBV9 (chr7:142239536-142240057). Pruning was then performed using the ‘--indep-pairwise’ option in PLINK with a window size of 1000 kilobases and a pairwise r2 threshold of 0.1, resulting in 201,017 sites that were then used to calculate the first 20 principal components (PCs) using PLINK, of which the top 10 were included as covariates in all *cis*-eQTL analyses.

#### Additional sample and variant filtering

The following additional filters were applied to the QC2 variant call set for *cis*-eQTL analyses: the 361 individuals (201 PM and 160 LIV) from the 445 that had WGS data in QC2 and also were retained for analysis following completion of all RNA sequencing quality control procedures were retained. After subsetting for these individuals, SNPs were removed for missingness rate greater than 2%, Hardy-Weinberg p-value < 5 x 10^−16^, and minor allele count less than 10. This left 5,136,250 SNPs for analysis. For protein *cis*-eQTL analyses, the subset of the 361 that also had LC-MS data were retained for analysis (N=345), but SNP filters were applied on the version of the dataset with 361 individuals.

### Main *cis*-eQTL analyses

For primary RNA, mature RNA, and protein data, the peer package (v1.0) was used in R (v4.0.3) to identify latent surrogate variables (SVs) independent of the variables used in the corresponding LIV-PM DE model (excluding LIV-PM variable itself and Individual ID). PEER was run 30 times, varying the number of SVs to identify from 1 to 30. After each PEER run, the SVs that were generated and the covariates used in DE with the exception of LIV-PM status and IID were regressed out of the expression data using the lmFit() function of the limma R package (v3.46.0) and the residuals() function of the base stats R package. The “cis” function of the QTLtools (v1.1) unix package was then run on the resulting 30 expression matrices for primary RNA, mature RNA, and protein expression after defining gene transcription start sites using the reference. The top 10 genetic ancestry PCs were included as covariates using the “--cov” parameter and the “--permute” parameter was set to 10,000 and “--window” set to the default of 1 megabase. The p-value of association between genotype and expression given by the fitted beta distribution and then adjusted for the number of variants tested in *cis* was further corrected for the number features tested using the qvalue() function of the qvalue R package (v2.22.0). After p-value adjustment, the number of *cis*-eQTL features was counted for the 30 different SV thresholds, and the threshold that produced the highest number of *cis*-eQTL features was selected as the number of SVs to account for in *cis*-eQTL analyses for primary RNA (30 SVs), mature RNA (28 SVs) and protein (30 SVs). The unadjusted p-values were the p-value of association between genotype and expression given by the fitted beta distribution and then adjusted for the number of variants tested in *cis*, but not adjusted for the number of expression features tested.

### Variance in expression explained by *cis*-eQTLs

To calculate the variance explained by *cis*-eQTL, the set of *cis*-eQTL identified by QTLtools at the optimal SVs were extracted. The genetic data was then subset for these and converted into a tab-delimited genotype matrix using plink (v1.90b6.21). This matrix was then read into R (v4.0.3) along with the voom-normalized form of the expression matrix for RNA or the normalized imputed data for protein. For each genotype-RNA/protein feature combination, the variance in the expression explained by the genotype was calculated using the fitExtractVarPartModel() function of the variancePartition R package (v1.20.0) and accounting for the covariates used in the corresponding LIV-PM DE model except for Individual ID. Then, the same procedure was carried out to calculate the variance in expression explained by LIV-PM status, except not including genotype in the formula. The Spearman’s correlation between the variances explained by *cis*-eQTL genotypes and the variances explained by LIV-PM status for RNA and protein features was calculated using the cor.test() function of the base stats R package. The overlap between DE features and *cis*-eQTL features was tested using the fisher.test() function of the base stats R package. The Fisher’s tests were performed using an iterative procedure where each iteration calculated the overlap between LIV-PM DE features and features associated with a *cis*-eQTL (i.e., CRFs) using increasingly stringent criteria to define CRFs. The CRF definitions were made more stringent with each iteration by increasing the minimum variance in feature expression required to be explained by the *cis*-eQTL for the feature to be defined as a CRF. The starting threshold was a VE >= 0, then the threshold was increased by 0.1 until there were less than 5 features remaining above the threshold. The set of p-values were adjusted using the FDR in p.adjust() and adjustment was done separately for each omic and VE (ie, the set of tests for primary RNA and VE of eQTL were adjusted for together but not with the set of tests for primary RNA and VE of DEG).

### Context-dependent *cis*-eQTL analyses

To test the hypothesis that the influence of *cis*-eQTL on RNA and protein feature expression differ between LIV samples and PM samples, regression models were run testing the association between feature expression and the interaction between *cis*-eQTL genotypes and LIV-PM status. Conceptually, this analysis examined if the slopes of the relationships between *cis*-eQTL genotypes and feature expression in LIV samples differed from the slopes of the relationships between *cis*-eQTL genotypes and feature expression in PM samples. Features where differences in these slopes were observed are referred to as CD-CRFs. The interaction QTL analysis was run using fastqtl (v2.184) with “--permute 100” and “--interaction” set to a path containing a file with one row per individual and two columns: one column for individual ID and a second column set either to 0 (LIV) or 1 (PM). The top 10 genetic PCs were included as covariates using the “--cov” parameter. The expression data input was the same expression data selected for *cis*-eQTL analyses after identifying the optimal number of SVs to regress out (so SVs and DE covariates had been removed). To ensure any interaction effects were not due to loci with different frequencies in LIV samples and PM samples, a GWAS was performed on LIV-PM status in the version of the genetic data used as input to the *cis*-eQTL analyses for RNA (N=361 individuals and 5,136,250 SNPs) using plink (v1.90b6.21) and including as covariates the first 20 genetic PCs using the “--logistic” command. Only interaction QTLs that had a unadjusted GWAS p-value > 0.05 were considered for analysis. Additionally, a frequency filter was applied such that every QTL had to have greater than 20 allele counts in LIV samples and greater than 20 allele counts in PM samples. After applying these filters, there were 21,008 primary RNA transcripts, 27,567 mature RNA transcripts, and 5,125 proteins in the context-dependent *cis*-eQTL analyses. To adjust the interaction QTL p-values the same approach was used as for *cis*-eQTLs. The unadjusted p-values were the p-value of association between genotype and expression given by the fitted beta distribution and then adjusted for the number of variants tested in *cis*, but not adjusted for the number of expression features tested. K-S tests were run using the ks.test() function of the base stats R package on the unadjusted p-values.

To compare CRFs identified in the current study (“LBP CRFs”) to postmortem brain CRFs from the published literature, summary statistics were downloaded from Supplementary Table 4 of a report by the AMPAD consortium^11^. PM CRFs were defined as genes in the sheets titled “eGenes” and “eProteins” where the column titled “FDR (BH)” had a value less than 0.05. LBP RNA CRFs were compared to PM RNA CRFs using a Fisher’s exact test implemented using the fisher.test() function of the base stats R package. The set of RNA transcripts included in the Fisher’s exact test was the set of RNA transcripts expressed in both the LBP and AMPAD samples. LBP Protein CRFs were compared to PM Protein CRFs using a Fisher’s exact test implemented using the fisher.test() function of the base stats R package. The set of proteins included in the Fisher’s exact test was the set of proteins expressed in both the LBP and AMPAD samples.

An example of one primary RNA, one mature RNA, and one protein that displayed significant associations with the genotype/LIV-PM interaction term are presented in ***Figure 3B*** (***Bottom Row***) - for these the data plotted is the expression after regressing out SVs and DE model covariates (but not genetic PCs).

### Rare PTV analyses in bulk RNA-seq and bulk LC-MS data

Variants in the QC2 callset were annotated for their predicted effect on protein function using the Variant Effect Predictor (VEP; v96) software with the LOFTEE plugin. PTVs were defined as any variant annotated by the LOFTEE plugin as “loss-of-function” with either “high confidence” or “low confidence”. Synonymous variants were those annotated by VEP as such in the “consequence” field of the VEP output (https://useast.ensembl.org/info/genome/variation/prediction/predicted_data.html). Population allele frequency annotations for each QC2 variant were obtained from the Genome Aggregation Database (gnomAD; v3.1)^35^. Allele frequencies in the LBP cohort were calculated on the set of individuals in QC2 (N = 445) using plink. Rare was defined as an alternate allele fraction less than 0.002 in LBP (which for 900 chromosomes equates to singletons) and less than 0.0001 in gnomAD.

As with the *cis*-eQTL analyses, one sample in the RNA and protein data was retained per individual for rare variant analyses. The same selection process for the *cis*-eQTL analyses was used (ie, the same samples) totaling 361 individuals in the RNA rare variant analyses (160 LIV and 201 PM) and 345 in the protein rare variant analyses (150 LIV and 195 PM).

To assess the effect of rare PTVs on RNA and protein expression levels LIV and PM samples were analyzed separately. The RNA transcripts and proteins included in the rare PTV analyses were limited to those where at least one individual in the dataset being analyzed (i.e., LIV samples or PM samples) had a variant in the QC2 callset that was annotated to that gene as either a rare PTV or a rare synonymous variant. This resulted in the following number of RNA transcripts and proteins analyzed: rare PTVs in primary RNA transcripts for LIV samples = 578; rare PTVs in primary RNA transcripts for PM samples = 552; rare PTVs in mature RNA transcripts for LIV samples = 652; rare PTVs in mature RNA transcripts for PM samples = 613; rare PTVs in protein for LIV samples = 166; rare PTVs in protein for PM samples = 169; rare synonymous variants in primary RNA transcripts for LIV = 4,904; rare synonymous variants in primary RNA transcripts for PM samples = 5004; rare synonymous variants in mature RNA transcripts for LIV samples = 5,325; rare synonymous variants in mature RNA transcripts for PM samples = 5,464; rare synonymous variants in protein for LIV samples = 1,724; rare synonymous variants in protein for PM samples = 1,845.

A transformation procedure was carried out on each RNA transcript and protein. RNA transcript and protein expression levels (using the residuals calculated on the full cohort) were transformed into ranks. The rank was simply the rank of sample S for feature F within the subset of the cohort being analyzed (e.g., LIV samples), such that resulting values ranged from one (i.e., sample S had the highest expression level of gene G) to the number of samples in the cohort being analyzed (i.e., sample S had the lowest expression of gene G). The ranks were then normalized by dividing the rank by the number of samples in the cohort being analyzed.

After the transformations were complete, the effect of rare variants on RNA and protein expression levels were assessed using the one-sided K-S test implemented through the ks.test() function of the base R stats package, with the alternative hypothesis specifying the expectation that carriers would have higher ranks (i.e., lower RNA transcript and protein expression levels) compared to non-carriers. For primary RNA, mature RNA and protein a total of four K-S tests were performed (LIV rare PTVs, PM rare PTVs, LIV rare synonymous variants, PM rare synonymous variants), totaling 12 tests. For each test, the ranks of carriers were compared to the ranks of non-carriers. As an example, consider the test for the effect of rare PTVs on primary RNA levels in LIV samples. As noted above, a LIV rare PTV carrier was identified for 578 primary RNA features, some of which had more than one LIV rare PTV carrier such that 610 total rare PTVs were seen in LIV for these 578 primary RNA features. The corresponding 610 ranks formed one set of values for the K-S test. The second set of values was comprised of the ranks of the noncarriers for the 578 primary RNA features (which formed a uniform distribution from 0-1). Since a total of 12 tests were performed, the significance of these K-S tests was assessed after performing a Bonferroni correction for 12 tests using the p.adjust() function of the base R stats package.

### Confounding effect of LIV-PM status on rare PTV effects

The relative effect of carrier status on mature RNA levels was compared to the effect of LIV-PM status on mature RNA levels. The following procedure was performed to do this:

1. Mature RNA LIV-PM DE signature was read into R
2. The mature RNA features for analysis were the 652 features with >=1 rare PTV carrier in LIV samples and the 613 features with >=1 rare PTV in PM samples. These covered 1214 unique features (some features had a LIV sample and a PM sample as a rare PTV carrier)
3. The DE status of the 1214 features was extracted from the DE results. The 652 mature RNA features with >=1 rare PTV in a LIV sample were 282 LIV DEGs, 189 PM DEGs, and 181 not DE. The 613 mature RNA features with >=1 rare PTV in a PM sample were 257 LIV DEGs, 190 PM DEGs, and 166 not DE.
4. The mature RNA residuals calculated on the full LBP cohort were used. For each of the 652 LIV features, the following procedure was performed:

a. LIV carriers were defined
b. LIV non-carriers were defined
c. PM non-carriers were defined
d. The LIV carriers were combined with the LIV non-carriers, and the ranking transformation was performed. The mean of the ranks was subtracted from the mean of the LIV carrier ranks, and this value was divided by the standard deviation of the ranks - this resulted in the Z-score for LIV carriers to LIV non-carriers.
e. The LIV carriers were combined with the PM non-carriers, and the ranking transformation was performed. The mean of the ranks was subtracted from the mean of the LIV carrier ranks, and this value was divided by the standard deviation of the ranks - this resulted in the Z-score for LIV carriers to PM non-carriers.
5. For each of the 613 PM features, the following procedure was performed:

a. PM carriers were defined
b. LIV non-carriers were defined
c. PM non-carriers were defined
d. The PM carriers were combined with the LIV non-carriers, and the ranking transformation was performed. The mean of the ranks was subtracted from the mean of the LIV carrier ranks, and this value was divided by the standard deviation of the ranks - this resulted in the Z-score for PM carriers to LIV non-carriers.
e. The PM carriers were combined with the PM non-carriers, and the ranking transformation was performed. The mean of the ranks was subtracted from the mean of the LIV carrier ranks, and this value was divided by the standard deviation of the ranks - this resulted in the Z-score for PM carriers to PM non-carriers.
6. To test the effect of PFC functional state on the rare PTV effects on mature RNA levels, the following procedure was performed:

a. The 652 features with a LIV carrier rare PTV were subset for the 282 LIV DEGs. A K-S test was performed using the base stats R package comparing the Z-scores for LIV carriers to LIV non-carriers to the Z-scores for LIV carriers to PM non-carriers.
b. The 652 features with a LIV carrier rare PTV were subset for the 189 PM DEGs. A K-S test was performed using the base stats R package comparing the Z-scores for LIV carriers to LIV non-carriers to the Z-scores for LIV carriers to PM non-carriers.
c. The 613 features with a PM carrier rare PTV were subset for the 257 LIV DEGs. A K-S test was performed using the base stats R package comparing the Z-scores for PM carriers to LIV non-carriers to the Z-scores for PM carriers to PM non-carriers.
d. The 613 features with a PM carrier rare PTV were subset for the 190 PM DEGs. A K-S test was performed using the base stats R package comparing the Z-scores for PM carriers to LIV non-carriers to the Z-scores for PM carriers to PM non-carriers.
e. Multiple test correction was performed for the four K-S tests using the method of Benjamini and Hochberg implemented in R using the p.adjust() function of the base stats package.

## Supplementary table descriptions

**Supplementary Table 1**

The FSCV DE signatures from snRNA-seq of 15 LIV samples

**Supplementary Table 2**

KEGG gene set enrichment results for FSCV DEFs in the FSCV DE signatures

**Supplementary Table 3**

The MER DE signatures from mature RNA STN and mature RNA GPi aperiodic exponents (115 LIV samples)

**Supplementary Table 4**

List of the 588 TPAWN genes

**Supplementary Table 5**

GWAS catalog gene set enrichment results for TPAWN genes

**Supplementary Table 6**

The LIV-PM DE signatures from the bulk RNA-seq and bulk LC-MS

**Supplementary Table 7**

KEGG gene set enrichment results for LIV DEGs and PM DEGs in the LIV-PM DE signatures.

**Supplementary Table 8**

KEGG enrichments in differentially correlated RNA-protein pairs

**Supplementary Table 9**

*cis*-eQTL results for full cohort (primary, mature, protein)

**Supplementary Table 10**

CD-*cis*-eQTL results for full cohort (primary, mature, protein)

